# Effect of maternal infection on stillbirths and early neonatal deaths: nested case-control studies in pregnancy cohorts in East Africa

**DOI:** 10.1101/2025.09.23.25335487

**Authors:** Anna C Seale, Meron Kebede, Rosanna Glazik, Mussie Brhane, Mulu Berihun, Zelalem Teklemariam, Dadi Marami, Joseph Oundo, Tadesse Gure, Tseyon Tesfaye, Ann Karanu, Gumbi Wilson, Christina W Obiero, Dianna M Blau, Helen Barsosio, Angela Koech, Samir Saha, Joy E. Lawn, Robert F Breiman, N Claire Gordon, Stefanie Wittmann, Lola Madrid, Abraham Aseffa, Yadeta Dessie, James A Berkley, Nega Assefa, J Anthony G Scott

## Abstract

**Background:** Reducing perinatal deaths is a priority, but data on maternal infectious causes are sparse in low- and middle-income countries where the burden is highest. We aimed to describe maternal infections at delivery and their association with perinatal death in Kilifi County Hospital (KCH), Kenya and Hiwot Fana Comprehensive Specialised Hospital (HFCSH), Ethiopia.

**Methods:** We investigated 642 mothers delivering stillbirths/newborns dying in the first 24h after birth (cases) and 855 mothers with newborns surviving ≥24h (controls), from well-characterised pregnancy cohorts in a nested case-control design in KCH (2011-17) retrospectively and HFCSH (2019-20) prospectively. We tested maternal blood for infection at delivery using molecular methods with 60 PCR targets (TaqMan Array Cards, TAC). In HFCSH, vagino-rectal swabs (VRS) and oropharyngeal swabs (OPS) were also tested, with 28 and 42 PCR targets respectively, along with conventional microbiological testing. We tested associations between maternal infection and perinatal death for each site, separately, and combined, and adjusted for potential confounders. We did a sensitivity analysis using only controls with good pregnancy outcomes in KCH. Where appropriate, we calculated the population attributable fraction (PAF).

**Results:** In HFCSH, maternal bacteraemia was associated with perinatal death (adjusted odds ratio aOR3.7 [1.5-9.2]). In KCH, bacterial detection in maternal blood was associated with perinatal death (aOR2.7 [1.2-6.0]), but only in sensitivity analysis. *Escherichia coli*/Shigella was associated with perinatal death when cultured/detected in blood (aOR2.6 [1.1-6.3]) across sites. Though infrequent, *Bordetella sp*. was associated with perinatal death (OR4.9[1.1-23.0]) on OPS in HFCSH. No other individual infections were associated with perinatal death. The PAF for perinatal deaths among hospital deliveries was 6.1% (4.0%-8.2%) for maternal bacteraemia in HFCSH and 4.9% (2.6%-7.2%) for bacterial detection in KCH.

**Conclusions:** Maternal bacterial infection is associated with perinatal death in high-burden settings, and in our study accounted for around 5% of perinatal deaths in hospital deliveries. The study was underpowered to detect species-specific infections associated with perinatal death.

**Funding:** Wellcome (205184)

## Background

Worldwide, deaths in children aged under five years have decreased from 12.7 million in 1990 to 4.9 million in 2022.^1^ However, neonatal deaths (days 0-27) have decreased more slowly, from 5.1 million to 2.3 million each year.^1^ A third of these deaths occur on the day of birth.^2^ In addition, there were an estimated 1.9 million stillbirths (babies delivered with no signs of life ≥28 weeks’ gestation) in 2021.^1^ Nearly a half of all neonatal deaths (1.1M in 2022) and stillbirths (0.9M in 2021) were in sub-Saharan Africa.^1^ Infection is thought to cause a quarter of all neonatal deaths^3^ and, although data are limited, accounts for approximately half of all stillbirths.^4^

Detailed data on infections in neonatal death, and especially of stillbirth, remain limited, but data on maternal infection associated with those deaths are even more scarce. The recent WHO Global Maternal Sepsis Study (GLOSS), highlighted the frequency of symptomatic maternal infection, especially in low- and middle-income countries,^5^ with systematic screening to ascertain clinically-defined cases of possible infection. However, the study was limited in that it only reported infectious aetiologies detected during routine investigation, rather than testing systematically for potential infections.^6^ Similarly, whilst multi-country trials of prophylactic azithromycin administration in pregnancy suggest improved outcomes,^7^ which specific infections are being treated remains unclear without systematic testing.

There are four ways in which maternal infections in pregnancy can affect the fetus. Firstly, haematogenous infections can spread across the placenta. This applies to *Treponema pallidum*,^8^ *Brucella sp*.,^9^ *Coxiella burnetii* (Q fever) and *Mycobacterium tuberculosis*.^*10*^ Among viruses, maternal cytomegalovirus^11^ and Zika infections have been reported in perinatal deaths,^12^ and hepatitis E may also contribute.^13^ HIV-1 infection contributes to adverse newborn outcomes, but its association with perinatal death is uncertain.^14^ Secondly, some maternal infections can result in placental insufficiency (e.g. malaria due to *Plasmodium falciparum*).^15^ Thirdly, pathogens causing infection of the maternal genitourinary tract may ascend to the uterus, for example *Escherichia coli* and group B streptococcus (GBS). These infections are leading bacterial causes of perinatal death,^16^ and are common causes of maternal sepsis in high-income countries (HICs) such as the UK.^17^ Fourthly, systemic disease in the mother, including bacterial sepsis or severe viral infections, such as influenza, dengue^18^ and chikungunya,^19^ are associated with fetal death through severe maternal illness which can, for example, compromise fetal-placental circulation.

Aetiologic studies in children have emphasised the importance of systematic case ascertainment and testing using both conventional microbiological methods (culture), despite low sensitivity,^20^ and molecular methods. Polymerase chain reaction (PCR) panels targeting bacterial and viral pathogens have provided important additional data in multi-country studies of gastroenteritis, pneumonia and neonatal sepsis.^21-23^ Learning from these studies, we systematically tested for maternal infection using both conventional and molecular microbiology, and determined their associations with perinatal death. We used a case-control study nested within well-described cohorts of pregnant women delivering in two settings in East Africa: Kilifi County Hospital, Kilifi, Kenya and Hiwot Fana Comprehensive Specialized Hospital, Harar, Ethiopia.

## Methods

### Study design and setting

In Kilifi County Hospital (KCH), a rural county hospital in coastal Kenya, with comprehensive obstetric care and 3-4000 deliveries per year, mothers were recruited at admission for delivery to an observational cohort study (2011 to 2017). Maternal data on antenatal history, delivery details, partogram, maternal problems up to hospital discharge, and newborn outcomes, including hospitalisation, were recorded by nurses in a structured maternity record. Maternal blood at admission was routinely collected and stored at −80°C.^24^ A similar prospective observational cohort study was established in Haramaya University Hiwot Fana Comprehensive Specialized Hospital (HFCSH, 2019-2020), a referral hospital in Harar, Eastern Ethiopia, with comprehensive obstetric care and 4-6000 deliveries per year. Here, maternal blood was collected and taken to Hararghe Health Research (HHR) laboratory for bacterial culture as well as storage, along with oropharyngeal swabs (OPS), and vagino-rectal swabs (VRS) using viral transport medium. The HHR laboratory also performed microbiological tests for Child Health and Mortality Prevention Surveillance (CHAMPS),^25^ which ran concurrently with this study, investigating child deaths using minimally invasive tissue sampling (post-mortem). The studies were separate but participants in one study could be recruited to participate in the other.

Within these two observational cohorts, we nested case-control studies to identify and test the association of different maternal infections with perinatal death. Cases were defined as mothers whose pregnancy yielded a stillbirth (≥28 weeks’ gestation or >1000grams when the gestational age was unknown) or very early neonatal death (<24hours following birth). Controls were mothers whose pregnancy yielded a liveborn child (≥28 weeks’ gestation or >1000grams, and surviving >24h after birth). Cases and controls were selected from their respective sampling frames using random number generation. Where samples had not been taken from those selected for study, additional participants were randomly selected to achieve the target sample size. Women with multiple deliveries were not excluded, but if any delivery met the case definition, they were included as a case. The study design is summarised in Supplementary Table 1 and adherence to STROBE case-control reporting guidelines is shown in Supplementary Table 2.^26^

### Clinical maternal and newborn data collection

Examination of mothers on admission included routine vital signs (respiratory rate, heart rate, temperature, blood pressure) as well as symphyseal-fundal height, cervical assessment and general examination. Maternal danger signs at triage suggesting need for emergency care included any of the following: airway not patent, respiratory rate >30 or <10 breaths/minute, systolic blood pressure >160 or <90 mmHg, diastolic blood pressure >90 mmHg, heart rate <40 or >120 beats/minute, unconscious or alert only to pain, and other obstetric emergencies (including imminent delivery) requiring immediate intervention. Maternal and newborn outcomes at delivery were also recorded, including sex, birthweight (using SECA digital scales). Babies who were admitted to neonatal units were followed up for survival to 24h.

### Microbiological tests

In KCH, total nucleic acid was extracted from maternal blood plasma and pellet samples (Roche High Pure Viral Nucleic Acid Kit, Roche, Switzerland) and stored at −80°C prior to PCR processing. In HHR laboratory, nucleic acid was extracted from maternal blood samples, OPS and VRS using Qiagen EZ1 Advanced XL Automated Nucleic Acid Purification System (QIAGEN/ Japan). In both sites, detection of pathogens from the extracted nucleic acid was done using TaqMan array cards (TAC) (Applied Biosystems/USA) ^27^ by amplifying using QuantStudio 7 Flex Real-Time PCR System (Applied Biosystems/Singapore). Analysis was done with CDC QuantStudio software; QS7 v1.2 CDC. Different TAC cards were used for each specimen type (targets detailed in Supplementary Table 3).

In the prospective study in HFCSH, we used conventional microbiological methods to detect maternal bacteraemia and test vagino-rectal swabs. For blood cultures, BACTEC (Becton Dickinson, South Africa) bottles were incubated at 37±2ºC in BACT/ALERT^®^ 3D Microbial Detection Systems (BioMérieux/USA) for five days; bottles alerting positive were Gram-stained and for Gram positive cocci sub-cultured on to Blood Agar (BAP), Trypticase soy agar (TSA) and MacConkey (MAC); Gram positive cells (yeast cells) were subcultured onto Sabouraud Dextrose Agar (SDA). For Gram negative rods/coccobacilli these were subcultured onto BAP, chocolate blood agar (CBA) and MAC, and for Gram negative diplococci onto TSA, BAP, and CBA. Frozen, stored vagino-rectal swabs were thawed and subcultured on BAP, CBA, MAC, GC agar, CHROMagar, and Xylose Lysine Deoxycholate agar. For BAP, CBA and MAC incubation was in 5% CO_2_; all the other media were incubated aerobically for 48h. Pathogens were identified by morphological appearance, Gram staining and standard biochemical tests (BioMérieux’s Analytical Profile Index, API 20 E, API 20 NE, API NH and API Staph).

### Sample size and statistical methods

We aimed to identify pathogens strongly associated (OR>3) with perinatal death where the prevalence of infection in controls was low (∼1%) (Supplementary Table 4). We increased the proportion of controls to cases in Kenya to enable a sensitivity analysis that included only controls with good pregnancy outcomes; those whose babies were born by a spontaneous vaginal delivery (cephalic), had a birthweight ≥2500g, and did not require admission to the neonatal unit. We calculated the odds ratios detected (2 to 8) based on varying prevalence of exposure in controls (0.001 to 0.1) and selected a sample size of 350 cases in each site, 350 controls in HFCSH, 500 controls in KCH.

We described maternal participants in the study from each site, in terms of socio-demographics (age, marital status, education level, nulliparity), and signs and symptoms of infection. These included history of fever, prolonged rupture of membranes >18h (PROM>18h), dysuria, positive urine dipstick test for nitrite and/or leucocyte, and emergency signs (defined above). All statistical analyses were conducted in Stata release 14 (StataCorp, College Station, TX). We tested the association of each maternal characteristic with perinatal death using logistic regression. We included maternal age in the final model as an *a priori* confounder. We used a similar approach for the characteristics of the participants’ babies, where we described birth weight and sex, and used logistic regression to test the association with stillbirth or early neonatal death. We included the sex of the baby in the final model as an *a priori* confounder. For both models we tested characteristics (p<0.1) in a multivariable model and down-selected based on p values (p<0.1) for the final model. For maternal participants and their babies, we did these analyses for each site separately, and for both sites combined, setting site (KCH/HFCSH) as a random effect. We repeated these analyses using multiple imputation to assess impact of missing data. We also did an exploratory analysis to test the association of signs and symptoms of infection with maternal bacteraemia.

For infections in maternal blood, we described TAC detections for bacteria, viruses, parasite and fungi and blood culture results for clinically significant maternal bacteraemia; we excluded cultures of likely contaminants (coagulase negative staphylococcus and Bacillus spp.) from this definition, but included these participants in analysis. We tested the association of clinically significant maternal bacteraemia with perinatal death using logistic regression; we also tested the associations of individual pathogens with perinatal death. We similarly tested the association of all bacteria detected by TAC, all viruses detected by TAC, and each individual target (bacteria, virus, parasite or fungus) detected by TAC, with perinatal death. We did this for each site separately and for both sites combined, specifying site as a random effect. We also tested the association of the combination of bacterial targets detected either with TAC or blood culture with perinatal death; we did this for both sites combined, specifying site as a random effect. Where numbers of detections, or bacterial isolates were small, we used Fisher’s exact test to assess associations. We did a sensitivity analysis using data from KCH alone, testing the association of TAC detections with perinatal death using a sub-group of controls with good pregnancy outcomes (see definition above), in case inclusion, in the control group, of participants with clinically significant, but non-fatal, infection biased our primary analyses towards the null.

For infections on maternal oropharyngeal swabs (OPS), and maternal vagino-rectal swabs (VRS), we described the TAC detections. We also described conventional microbiological culture results from VRS. We tested the association of each target detected with perinatal death and each positive culture with perinatal death using logistic regression.

For results where infections were associated with perinatal death (p<0.1) we used multivariable models to adjust for potential confounders (maternal age, education level, marital status, nulliparity, baby/fetal sex) and down-selected based on p values (p<0.1) for the final model, including maternal age and baby/fetal sex as *a priori* confounders. We repeated these analyses using multiple imputation to assess impact of missing data.

We calculated the population attributable fraction (PAF) for results where infections were associated with perinatal death and there was good evidence for causality, based on assessment using the Bradford-Hill criteria. PAFs were calculated using the punafcc command in Stata release 14, based on maximum likelihood estimation of the attributable fraction from multivariable logistic models.^28^

We identified cases which were also included in CHAMPS,^25^ to compare findings from maternal investigation in this study and stillbirth/neonatal post-mortem investigations in that study that were used to determine cause of death.

### Patient and public involvement in research

Patients and the public were not involved in setting the research question or outcome measures, but they were involved in the design and conduct of the study through community engagement meetings.

## Ethics

This study was approved by the London School of Hygiene & Tropical Medicine, London, UK (14381); Kenya Medical Research Institute (KEMRI/SERU/CGMR-C/089/3479); Institutional Health Research Ethics Review Committee (IHRERC) College of Health and Medical Sciences, Harar Campus, Haramaya University (IHRERC/143/2017); Armauer Hansen Research Institute (AHRI/ALERT Ethics Review Committee (AAERC Protocol No. PO33/17)) and the Ethiopian Ministry of Science and Technology (310/298/2017).

## Results

### Participants

In KCH 25,414 pregnant women attended the labour ward between 1 Jan 2011 and 5 June 2017. In HFCSH, 3204 pregnant women attended the labour ward between 22 May 2019 and 5 September 2020. Following randomisation, in the final analysis, there were 318 cases and 508 controls from KCH and 324 cases and 347 controls from HFCSH (Figure 1). Among 318 cases in KCH, 269 (85%) had a stillbirth and 49 (15%) had a newborn death (<24h). Among 324 cases in HFCSH, 256 (79%) had a stillbirth and 69 (21%) had a newborn death (<24h).

**Figure 1:**
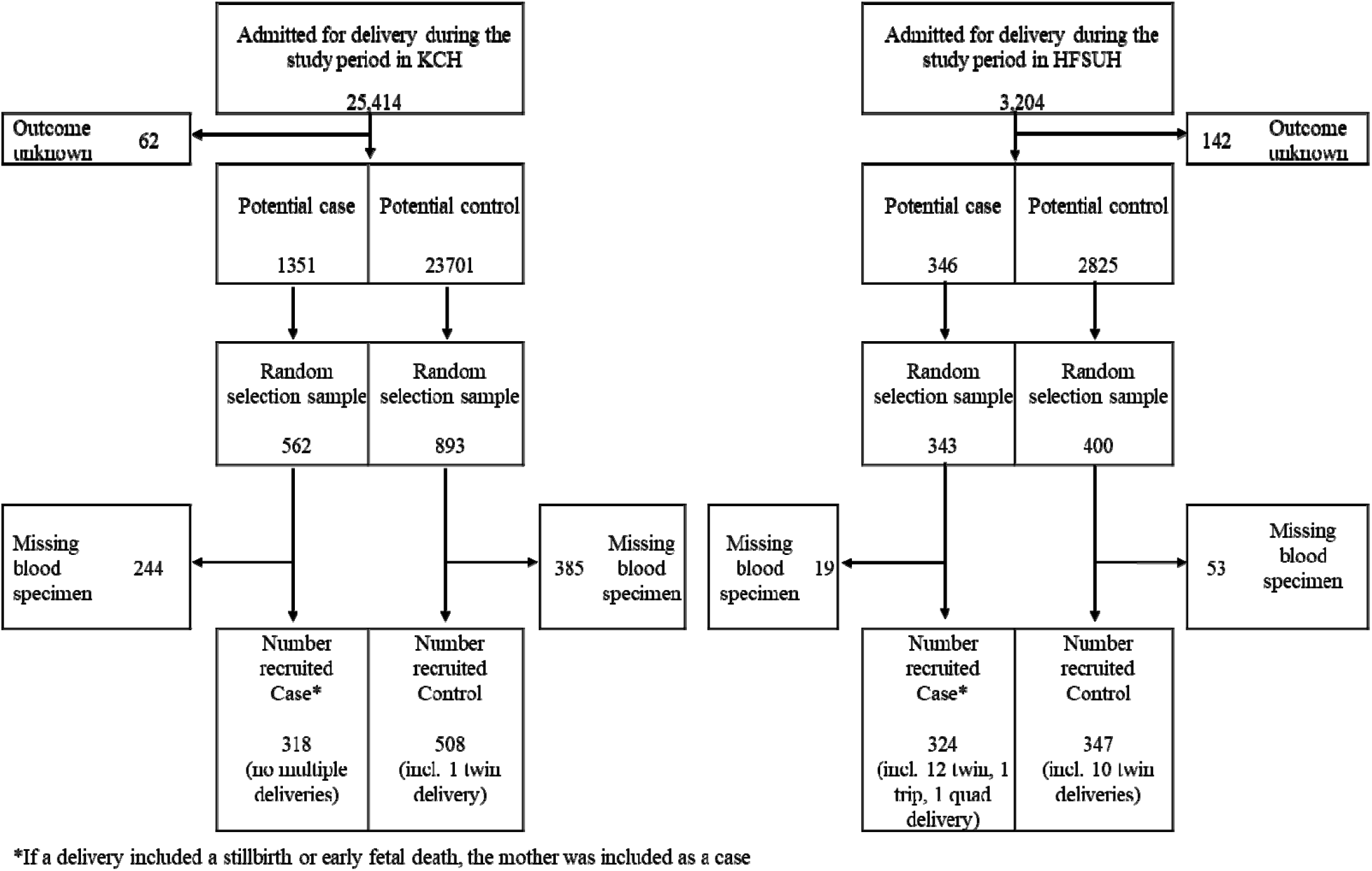
Recruitment of cases and controls from among pregnant women delivering in Kilifi County Hospital, Kenya (1 Jan 2011-5 Jun 2017) and Hiwot Fana Comprenhensive Specialised Hospital, Ethiopia (22 May 2019-5 Sep 2020).

In both KCH and HFCSH, a large majority of the women were married and multiparous (Table 1). The majority of women were educated to primary level in KCH (59% of participants in the control group), but not in HFCSH (26% of controls). Signs and symptoms of infection (described above) were infrequent, but women commonly presented with a sign of an emergency (89% and 49% of participants in control group, for KCH and HFCSH respectively). In the combined analysis across sites, socio-demographic and clinical signs associated with perinatal death included: no education (adjusted odds ratio aOR1.9 [1.5-2.5]), and emergency signs at delivery (aOR1.9[1.5-2.4]). Among the characteristics of babies, low and very low birthweight were associated with perinatal death (aOR3.7 [2.8-4.9] and aOR18.3 [10.2-32.5], respectively). Site-specific analyses are detailed in Supplementary Table 5 and 6, combined in Supplementary Table 7. Estimates of effect (odds ratios) for characteristics associated with perinatal death were comparable when multivariable analyses were repeated with imputation (data not shown). In the exploratory analysis, a history of fever (aOR6.1 [95% CI: 1.1-33.0]) and emergency signs at delivery (OR2.1[1.1-5.1]) were associated with maternal bacteraemia (Supplementary Table 8) in HFCSH.

**Table 1:** Characteristics of mothers and babies included in the study, by case and control status, for Kilifi County Hospital, Kenya and Hiwot Fana Comprehensive Specialised Hospital, Ethiopia.

| Maternal characteristic | Kilifi County Hospital |  |  |  | Hiwot Fana Comprehensive Specialised Hospital |  |  |  |
| --- | --- | --- | --- | --- | --- | --- | --- | --- |
|  | Cases<br>N=318 | (%) | Controls<br>N=508 | (%) | Cases<br>N=324 | (%) | Controls<br>N=347 | (%) |
| **Age, years** |  |  |  |  |  |  |  |  |
| <20 | 43 | 13.5 | 96 | 18.9 | 33 | 10.2 | 30 | 8.6 |
| 20 to <30 | 157 | 49.4 | 277 | 54.5 | 165 | 50.9 | 218 | 62.8 |
| 30 to <40 | 98 | 30.8 | 124 | 24.24 | 97 | 29.9 | 68 | 19.6 |
| ≥ 40years | 15 | 4.7 | 10 | 2.0 | 5 | 1.5 | 10 | 2.9 |
| Missing | 24 | 7.5 | 21 | 4.1 | 24 | 7.4 | 21 | 6.1 |
| **Marital status** |  |  |  |  |  |  |  |  |
| Married | 278 | 87.4 | 457 | 90.0 | 305 | 94.1 | 321 | 92.5 |
| Single | 26 | 8.2 | 43 | 8.5 | 1 | 0.3 | 1 | 0.3 |
| Divorced | 3 | 0.9 | 2 | 0.4 | 0 | 0.0 | 0 | 0.0 |
| Widowed | 0 | 0.0 | 0 | 0.0 | 0 | 0.0 | 0 | 0.0 |
| Missing | 11 | 3.5 | 6 | 1.2 | 18 | 5.6 | 25 | 7.2 |
| **Education level** |  |  |  |  |  |  |  |  |
| None | 64 | 20.1 | 73 | 14.4 | 219 | 67.6 | 135 | 38.9 |
| Primary | 181 | 56.9 | 299 | 58.9 | 38 | 11.7 | 91 | 26.2 |
| Secondary | 55 | 17.3 | 88 | 17.3 | 16 | 4.9 | 51 | 14.7 |
| Higher | 7 | 2.2 | 33 | 6.5 | 24 | 7.4 | 36 | 10.4 |
| Missing | 11 | 3.5 | 15 | 3.0 | 27 | 8.3 | 34 | 9.8 |
| **Nulliparous** |  |  |  |  |  |  |  |  |
| No | 209 | 65.7 | 322 | 63.4 | 205 | 63.3 | 219 | 63.1 |
| Yes | 102 | 32.1 | 174 | 34.3 | 90 | 27.8 | 100 | 28.8 |
| Missing | 7 | 2.2 | 12 | 2.4 | 29 | 9.0 | 28 | 8.1 |
| **History of fever** |  |  |  |  |  |  |  |  |
| No | 305 | 95.9 | 498 | 98.0 | 290 | 89.5 | 320 | 92.2 |
| Yes | 8 | 2.5 | 5 | 1.0 | 3 | 0.9 | 4 | 1.2 |
| Missing | 5 | 1.6 | 5 | 1.0 | 31 | 9.6 | 23 | 6.6 |
| **Prelabour Rupture of Membranes >18h** |  |  |  |  |  |  |  |  |
| No | 268 | 84.3 | 438 | 86.2 | 256 | 79.0 | 271 | 78.1 |
| Yes | 15 | 4.7 | 24 | 4.7 | 38 | 11.7 | 44 | 12.7 |
| Missing | 35 | 11.0 | 46 | 9.1 | 30 | 9.3 | 32 | 9.2 |
| **Dysuria** |  |  |  |  |  |  |  |  |
| No | 309 | 97.2 | 488 | 96.1 | 291 | 89.8 | 323 | 93.1 |
| Yes | 3 | 0.9 | 15 | 3.0 | 1 | 0.3 | 2 | 0.6 |
| Missing | 6 | 1.9 | 5 | 1.0 | 32 | 9.9 | 22 | 6.3 |
| **Positive nitrite and/or leucocyte** |  |  |  |  |  |  |  |  |
| No | 216 | 67.9 | 344 | 67.7 | 103 | 31.8 | 125 | 36.0 |
| Yes | 61 | 19.2 | 117 | 23.0 | 41 | 12.7 | 52 | 15.0 |
| Missing | 41 | 12.9 | 47 | 9.3 | 180 | 55.6 | 170 | 49.0 |
| **Emergency signs\*** |  |  |  |  |  |  |  |  |
| Yes | 231 | 72.6 | 451 | 88.8 | 111 | 34.3 | 170 | 49.0 |
| No | 87 | 27.4 | 57 | 11.2 | 213 | 65.7 | 177 | 51.0 |
| **Mode of delivery** |  |  |  |  |  |  |  |  |
| Vaginal | 249 | 78.3 | 400 | 78.7 | 250 | 77.2 | 257 | 74.1 |
| Caesarean section | 60 | 18.9 | 96 | 18.9 | 74 | 22.8 | 90 | 25.9 |
| Missing | 9 | 2.8 | 12 | 2.4 | 0 | 0.0 | 0 | 0.0 |

| **Baby characteristics** | **N =318** | **%** | **N =509** | **%** | **N= 338** | **%** | **N =356** | **%** |
| --- | --- | --- | --- | --- | --- | --- | --- | --- |
| **Birthweight (g)** |  |  |  |  |  |  |  |  |
| 1000-1500 | 56 | 17.6 | 8 | 1.6 | 58 | 17.2 | 7 | 2.0 |
| 1500- 2500 | 88 | 27.7 | 81 | 15.9 | 87 | 25.7 | 32 | 9.0 |
| Over 2500 | 147 | 46.2 | 412 | 80.9 | 142 | 42.0 | 291 | 81.7 |
| Missing | 27 | 8.5 | 8 | 1.6 | 51 | 15.1 | 26 | 7.3 |
| **Sex** |  |  |  |  |  |  |  |  |
| Male | 150 | 47.2 | 227 | 44.6 | 177 | 52.4 | 176 | 49.4 |
| Female | 144 | 45.3 | 271 | 53.2 | 124 | 36.7 | 151 | 42.4 |
| Missing | 24 | 7.5 | 11 | 2.2 | 37 | 10.9 | 29 | 8.1 |

\* Maternal danger signs included any of the following airway not patent, respiratory rate >30 or <10 breaths/minute, systolic blood pressure >160 or <90 mmHg, diastolic blood pressure >90 mmHg, heart rate <40 or >120 beats/minute, unconscious or alert only to pain, and other obstetric emergencies (including imminent delivery) requiring immediate intervention.

### Detection of infection and association with perinatal death/stillbirth

In HFCSH, maternal bacteraemia occurred in 24/324 (7.4%) cases and 10/347 (2.9%) controls (OR 2.7 [1.3-5.7], aOR3.7 [1.5-9.2], Supplementary Table 9). In KCH, in our sensitivity analysis including only mothers with good pregnancy outcomes as controls, bacterial detection in maternal blood with TAC was associated with perinatal death (24/318 (7.5%) cases, 10/315 (3.2%) controls, OR2.5 [1.2-5.3], aOR2.7 [1.2-6.0], Supplementary Table 10). We did not otherwise find evidence of an association between maternal blood bacterial detection by TAC and perinatal death at either site, or at both sites combined.

For specific pathogens (Figure 2A), only *Escherichia coli* bacteraemia was associated with perinatal death (Fisher’s exact test p=0.026). However, maternal bacteraemia with *Escherichia coli* (n=5), *Streptococcus agalactiae* (n=2), *Streptococcus pyogenes* (n=1), Streptococcus Group C&G (n=3), *Burkholderia cepacia* (n=3) and *Salmonella paratyphi* (n=1), whilst rare, occurred only in cases. In the combined analysis of maternal blood cultures and maternal blood bacterial DNA detection using TAC, *E. coli* culture and/or *E. coli*/*Shigella spp*. detection was associated with perinatal death across sites (18/642 (2.8%) cases, 10/855 (1.2%) controls OR 2.2[1.0-4.8], aOR2.6 [1.1-6.3], Supplementary Table 11). *Treponema pallidum* was not associated with perinatal death (OR0.7 [0.3-1.6], for both sites combined). Detailed results for maternal bacteraemia, maternal blood target detection and association with perinatal death for HFCSH, KCH and the two sites combined are in Supplementary Tables 12,13,14, and 15, respectively. Details of the sensitivity analysis results are in Supplementary Table 16, Supplementary Figure 1, and details of the maternal blood targets detected by PCR and/or cultured combined across both sites in Supplementary Table 17.

**Figure 2A:**
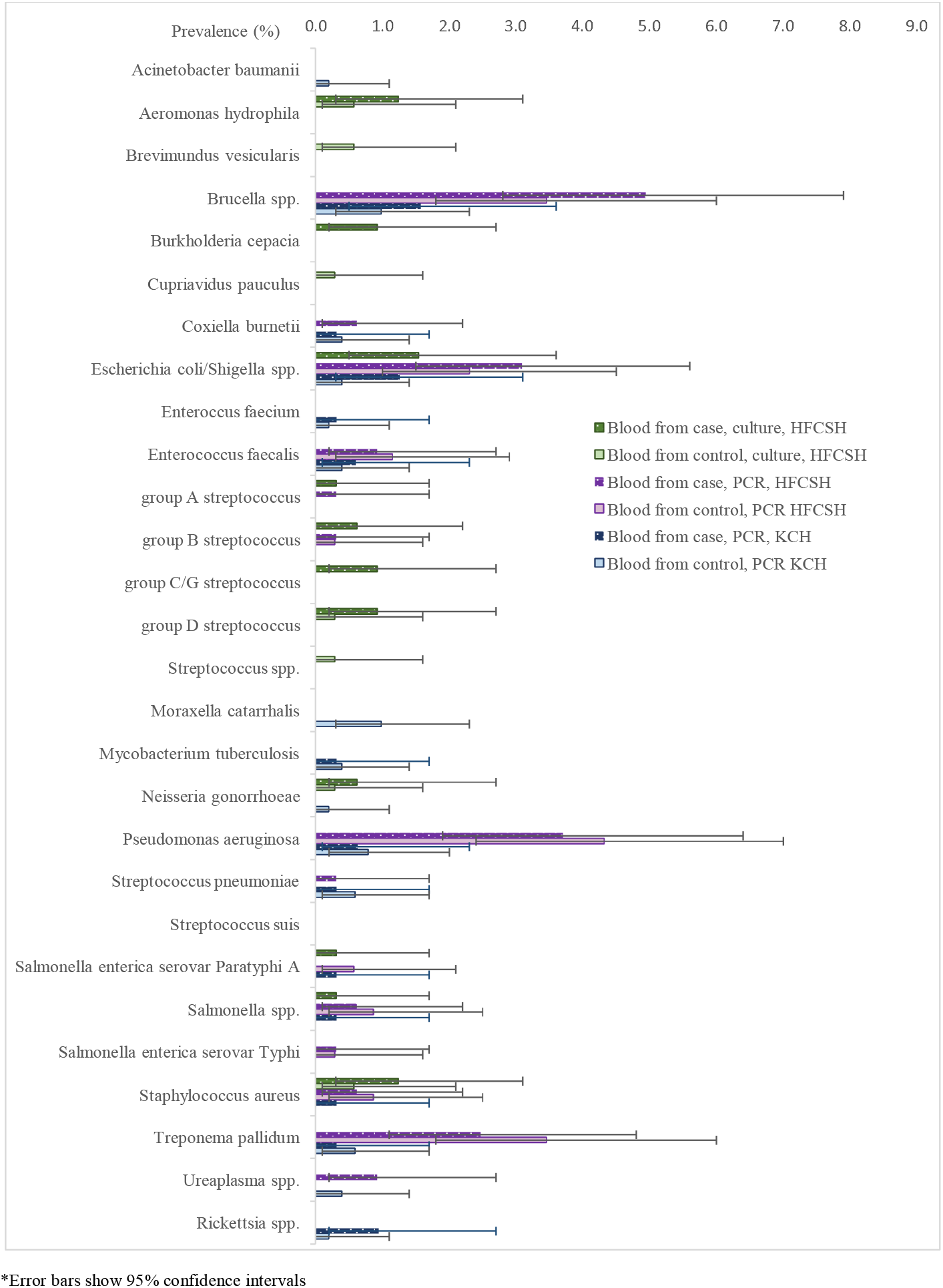
Prevalence of bacterial species, detected by PCR in blood in maternal participants in Kilifi County Hospital (KCH), Kenya, and prevalence of bacterial species detected by PCR or culture in blood, in maternal participants in Hiwot Fana Comprehensive Specialised Hospital (HFCSH), Ethiopia.* ^*^Error bars show 95% confidence intervals Figure 2B: Prevalence of viral, parasitic and fungal species, detected by PCR in blood in maternal participants in Kilifi County Hospital (KCH), Kenya and maternal participants in Hiwot Fana Comprehensive Specialised Hospital (HFCSH), Ethiopia.* ^*^Error bars show 95% confidence intervals

**Figure 2B:**
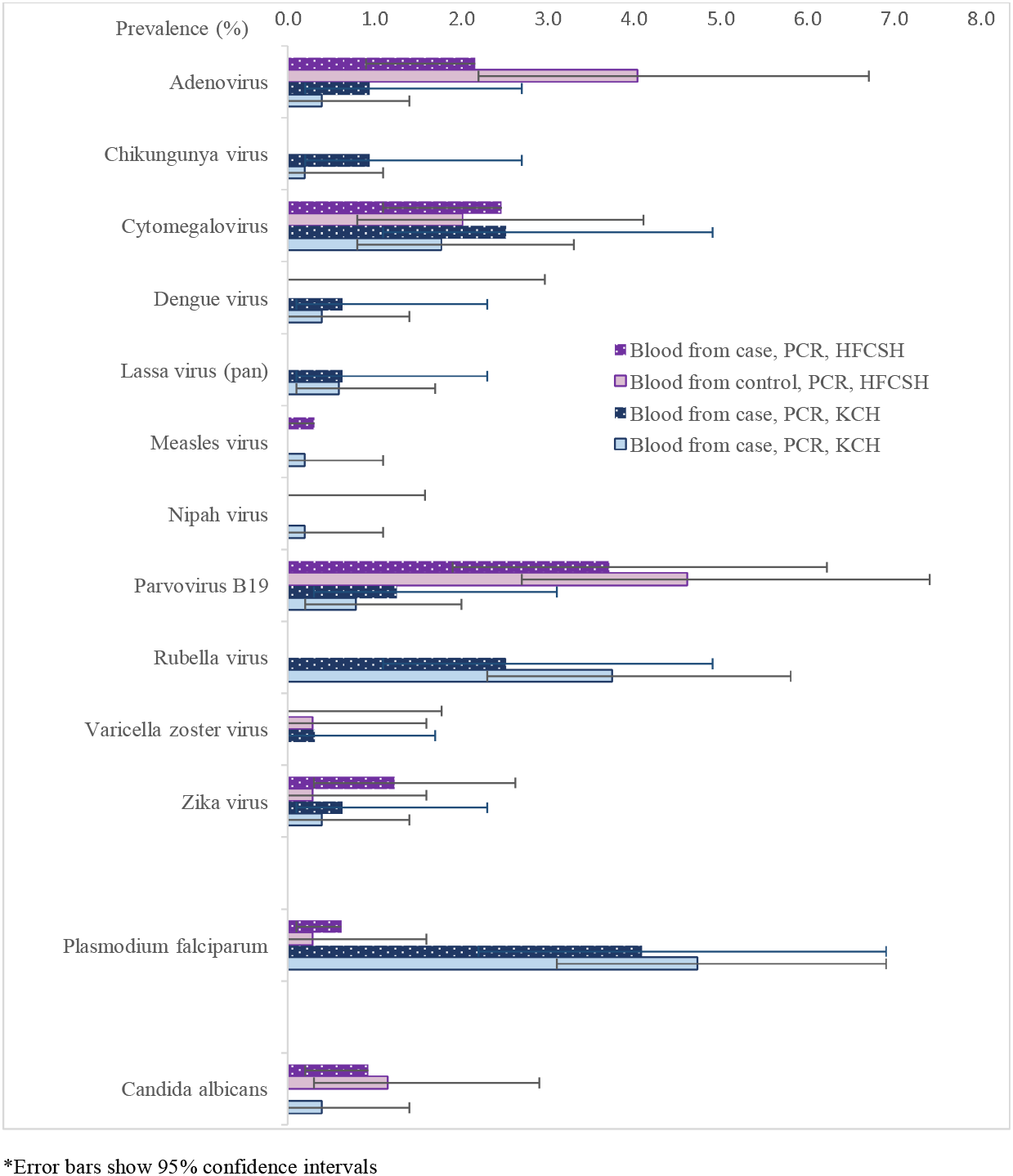
Prevalence of viral, parasitic and fungal species, detected by PCR in blood in maternal participants in Kilifi County Hospital (KCH), Kenya and maternal participants in Hiwot Fana Comprehensive Specialised Hospital (HFCSH), Ethiopia.* ^*^Error bars show 95% confidence intervals

We did not find evidence of an association between viral targets in maternal blood detected by TAC, individually or combined, and perinatal death. Viral targets commonly detected in maternal blood were cytomegalovirus and parvovirus B-19 in both sites (1.9% and 2.3% of participants in control group respectively), adenovirus in HFCSH (4.0% controls) and rubella at KCH (3.7% controls). Zika virus was detected, at low levels at both sites (0.4% controls) but other *Aedes sp*. mosquito-borne viruses, e.g. chikungunya and dengue viruses (0.2% and 0.4% controls, respectively), were only detected in KCH (Figure 2B, Supplementary Tables 13,14 and 15 for KCH, HFCSH, combined).

We did not find an association between molecular detection in maternal blood of any individual parasitic or fungal targets and perinatal death. The only parasite detected was *Plasmodium falciparum*, which was common in KCH (4.7% controls). The only fungus detected was *Candida albicans*, which was more common in HFCSH (2.4% controls). We did not find evidence of *P. falciparum* being associated with perinatal death (OR 0.9[0.5-1.8], Figure 2B, Supplementary Tables 13,14 and 15 for KCH, HFCSH, combined).

For maternal vaginal-rectal swab (VRS) samples, we found some evidence of an association between molecular detection of *E. coli/Shigella spp*. with perinatal death (140/324 (43%) cases, 111/347 (32%) controls OR[1.6; 1.2-2.2]; adjOR1.3 [0.9-2.9], Supplementary Tables 18, 20. However, we found no association between any individual conventional bacterial culture from VRS and perinatal death (Supplementary Table 19). Group D Streptococcus, commonly cultured (45% controls), was not included in the TAC panel (Figure 3).

**Figure 3:**
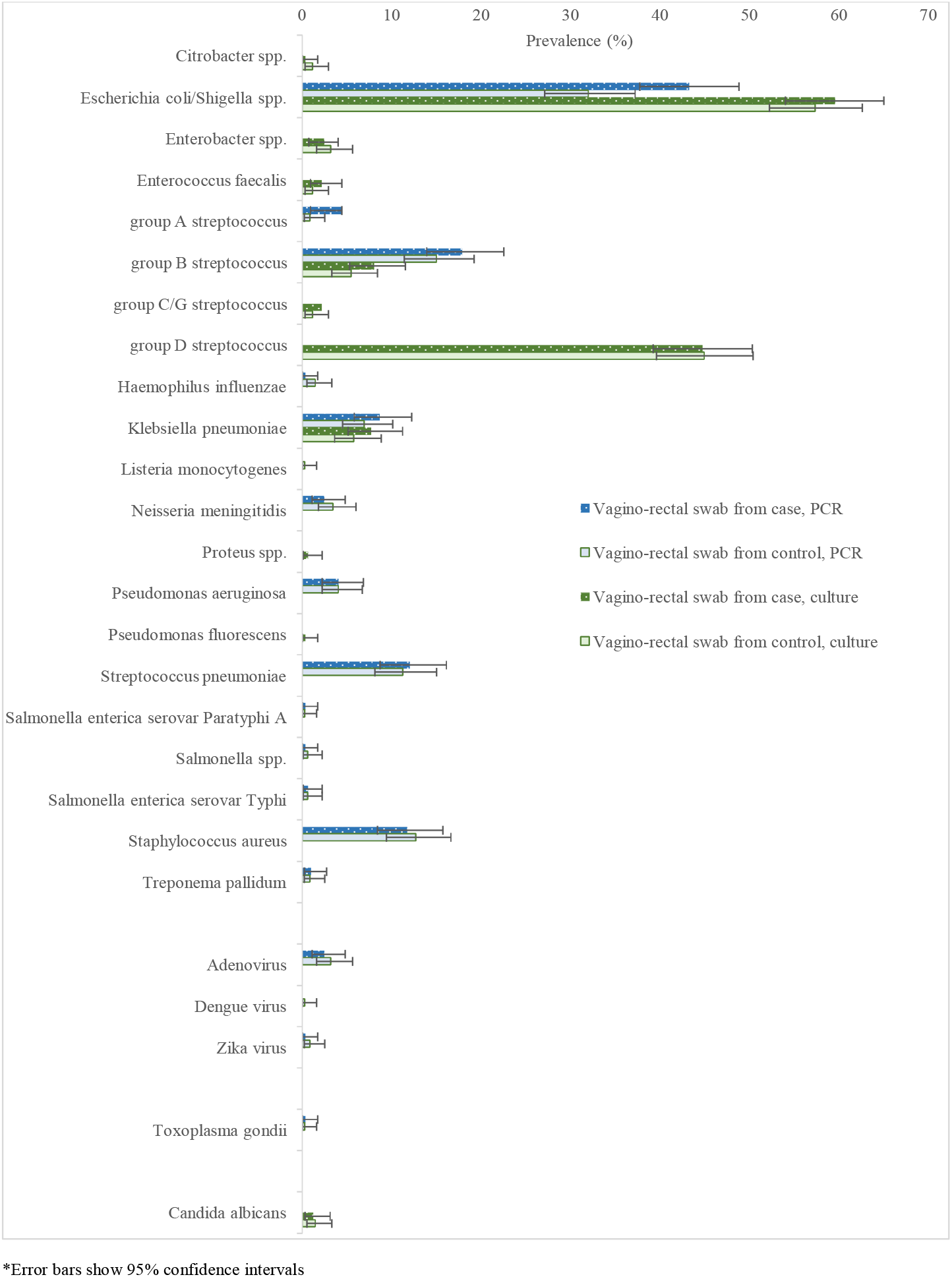
Prevalence of bacterial, viral, parastic or fungal species detected by PCR or bacterial culture from vagino-rectal swabs in maternal participants in Hiwot Fana Comprehensive Specialised Hospital, Ethiopia.* ^*^Error bars show 95% confidence intervals

For oropharyngeal swab (OPS) samples, we found that molecular detection of *Bordetella spp*. (pertussis or parapertussis) was associated with perinatal death (9/324 (2.8%) cases, 2/347 (0.6%) controls OR4.9[1.1-23.0]. Multivariable analyses only included Bordetella spp. in cases (missing data), so aORs are not reported. We did not find an association between molecular detection on OPS of any other individual bacterial, viral or fungal target and perinatal death (Supplementary Table 21, Supplementary Table 22). The bacterial target detected most frequently was *Streptococcus pneumoniae* (24.5% controls, Figure 4).

**Figure 4:**
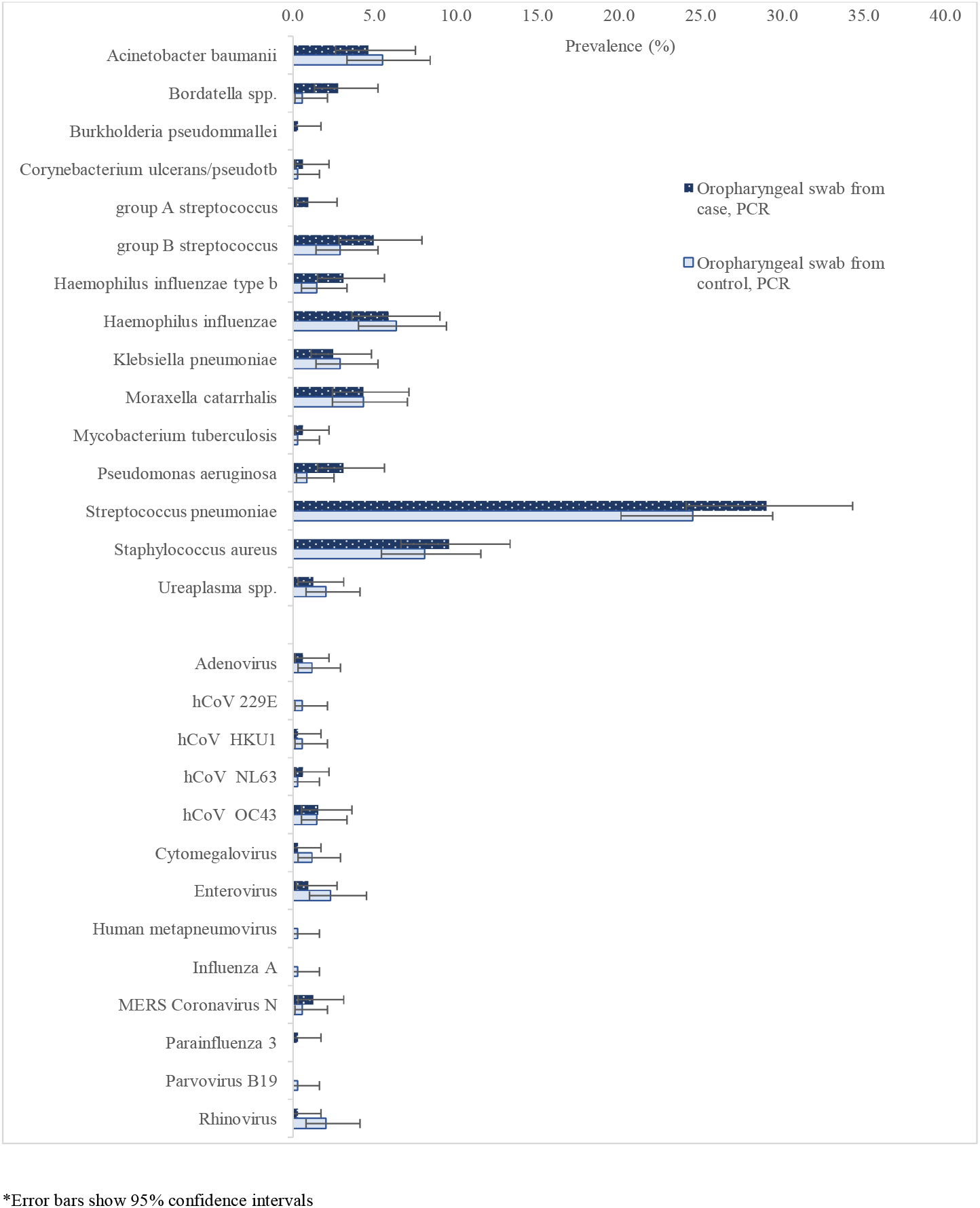
Prevalence of bacterial and viral species detected by PCR in oropharyngeal swabs from maternal participants in Hiwot Fana Comprehensive Specialised Hospital, Ethiopia.* ^*^Error bars show 95% confidence intervals

Estimates of effect (odds ratios) for infections associated with perinatal death were when repeated with multiple imputation, and comparable to the unadjusted estimate of effect for *Bordetella spp*. (data not shown).

### Estimates of the population attributable fraction (PAF)

Maternal bacteraemia in participants in HFCSH was associated with perinatal death. The sensitivity analysis of mothers with good pregnancy outcomes as controls in KCH was consistent with this result, which was specific to bacteria, and biologically plausible. The PAF of maternal bacteraemia, in participants in HFCSH, to perinatal death was 6.1% (4.0%-8.2%), and for maternal bacterial infection in participants in KCH (sensitivity analysis) was 4.9% (2.6%-7.2%).

### Comparison with Child Health and Mortality Preventions Surveillance (CHAMPS) data

We identified 14 cases where data were available on the cause of death from CHAMPS. ^25^ Of these, the underlying cause of stillbirth/early neonatal death was intrauterine hypoxia in 10/14, congenital abnormality in 2/14, intrauterine growth restriction in 1/14 and neonatal sepsis with *E. coli* in a preterm neonate in 1/14. In the neonatal sepsis case we did not detect *E. coli* bacteria in maternal blood (TAC or culture), but *E coli/Shigella* was detected using TAC from VRS, and *E. coli* was cultured from VRS. Conversely, there were two cases (one recorded as intrauterine hypoxia, one as intrauterine growth restriction) where maternal bacterial infection was cultured in blood (*Burkholderia cepacia*, and *Aeromonas hydrophila/caviae* respectively), and one case (intrauterine hypoxia) where maternal bacterial infection was detected using TAC in blood (*Pseudomonas aeruginosa)*.

## Discussion

Bacterial infections of mothers at delivery are associated with perinatal death in high-burden settings in East Africa. *Escherichia coli* is a probable contributor, but other infections are also likely to be important. In our study, around 5% of perinatal deaths in hospital were attributable to maternal bacterial infection at the time of delivery. We did not find evidence associating viral, parasitic or fungal infections at delivery with perinatal death.

Our study was strengthened through the inclusion of two sites with contrasting ecologies, allowing assessment of similarities and differences across sites, and it benefitted from testing multiple sample types, using both conventional and molecular methods to detect infection. However, these strengths also led to complexity and, during analysis, we undertook a high number of statistical tests. Where results are consistent, and there is a high level of plausibility, we have higher confidence in our findings, such as the overall association between bacterial blood infection and perinatal death. However, as an exploratory analysis, intended to highlight important infections for future study, we did not make a formal correction for the potential for chance, and with this limitation, we recognise that the evidence here is insufficient to confirm true associations for species-specific infections. There were, however, few specific infections (*E coli, Bordetella spp*.) where there was evidence of an association with perinatal death. The prevalence of individual infections was lower than we expected and whilst we would have detected strong associations (odds ratio >3) with a prevalence of exposure in the controls of around 1%, many infections in blood that we detected were less prevalent than this. In addition, whilst our study benefited from the sensitivity of molecular methods, the potential decrease in clinical specificity using molecular detection may have increased the probability of a null finding. Inclusion, in the control group, of participants with clinically significant, but non-fatal, infection may also have biased our primary analyses towards the null.

We found an association between culture of *E. coli* in maternal blood and perinatal death. Whilst recognising the potential role of chance here, it is biologically plausible with ascending infection around birth, and it is consistent with independent cord blood studies of neonatal sepsis at KCH.^29^ We also found maternal recto-vaginal colonisation with *E. coli* was associated with perinatal death. This may reflect ascending infection, but there was also evidence of confounding consistent with the observation that *E. coli* vagino-rectal colonisation is more common in lower socio-economic groups,^30^ who have an increased risk of perinatal death overall. Similarly, the association between *Bordetella spp*. and perinatal death may be true, but it may also be due to chance, or confounding; poor uptake of whole cell infant pertussis vaccination (and thus potential increased risk of maternal infection) is independently associated with poverty and/or lack of access to health care.

Some infections were sufficiently prevalent in controls (∼ 2% in blood), to provide the study with adequate power to detect a moderate association (odds ratios >2.5) with perinatal death. This applied to *Brucella spp*., *Pseudomonas aeruginosa* and *Treponema pallidum*, adenovirus, CMV, parvovirus B19, rubella and to *Plasmodium falciparum*. However we did not find good evidence of associations between these infections and perinatal mortality. For infections known to cause adverse perinatal outcomes in LMICs, which are the target of preventive programs, such as syphilis (caused by *Treponema pallidum*) and malaria (caused by *Plasmodium falciparum*) this lack of an association with perinatal death is potentially important, but does not exclude their contribution to other adverse perinatal outcomes, such as low birth weight, or congenital syphilis with attendant long-term sequelae. The time between infection and the perinatal outcome may also obscure a potentially causal association. Ascending (bacterial infections) such as *E. coli* or group B streptococcus are likely to act at, and around, delivery, whereas other infections (particularly viral) have a longer lag between infection and adverse perinatal outcome and would not be detected in our study design. CMV, Parvovirus B-19 and rubella, for example, have a higher risk of adverse outcome when infection takes place early in pregnancy. Another (non-viral) example in LMICs could be *Brucella spp*., as brucellosis more commonly causes early pregnancy loss before 24 weeks’ gestation; such cases would not have been included in our study.^31^ We also note the absence of *Klebsiella pneumoniae* bacterial infection in maternal blood, despite increasingly common reports of is contribution to neonatal sepsis and neonatal death in high burden settings.^16^ This reflects the differing pathogenesis of *Klebsiella pneumoniae*, acquired postnatally, frequently in preterm neonates admitted to hospital, compared to the ascending infection more frequent for *E. coli* or group B streptococcus infections.

Our study suggests around 5% of the perinatal deaths in the hospitals we studied were attributable to maternal bacterial infection. There was consistency across study sites, despite differences, for example, in maternal education levels and the range of infections across our sites, implying a degree of generalisability. These findings may not, however, be generalisable to lower levels of health care, or home deliveries; pregnant women delivering in hospitals are a biased sample, often with anticipated or emerging complications, and may have had intrapartum antibiotics. Whether the perinatal deaths we attribute to maternal bacterial infection are already being attributed to infection in the child, rather than the mother, is difficult to distinguish. In the 14 cases where we were able to compare investigations of both the mother and the baby there was no evidence of infection in the three perinatal deaths born to mothers with evidence of bacterial infection in blood. Conversely, one of the 14 babies experienced early neonatal death with evidence of *E. coli* infection, but we did not detect *E. coli* in maternal blood.

Our study of maternal infection covered a substantial number of targets on TAC, but there are other infections not included, that are common in these settings, for example *Borrelia burgdorferi* (lyme disease) is associated with adverse pregnancy outcomes in North America^32^ and *Borrelia duttoni* (causing tick-borne relapsing fever) is endemic in sub-Saharan Africa and has been linked to perinatal mortality in Tanzania.^33^ Little is known on the risk of *Borrelia recurrentis* (causing louse borne relapsing fever), which is endemic in Ethiopia. However, use of TAC can also improve sensitivity over conventional microbiology, particularly for pathogens, like group B streptococcus, that require an additional enrichment step in conventional culture.

Overall, our study indicates an important association between bacterial infections of maternal blood at delivery, and perinatal death. In hospital-based deliveries in East Africa, maternal bacterial blood infection may account for around 5% of perinatal deaths. Our study was underpowered for specific infections, and a much larger study would be required to test associations for specific infections with perinatal death. Ideally this would be a nested case-control study within a community setting, to reflect the population. Sampling maternal blood earlier in pregnancy may be needed to find true associations for infections which have the highest risk in the first, or early second, trimester of pregnancy, as well as at delivery. Improving our understanding of the aetiology of maternal infection associated with perinatal death in high-burden settings is important to inform targeted interventions and reduce the burden of death at birth.

## Data Availability

Data available on reasonable request and with institutional and ethical approvals.

## Acknowledgements

We would like to thank the Child Health and Mortality Prevention Surveillance (CHAMPS) team at Haraghe Health Research (HHR) Partnership for their collaboration with this work, as well as the CHAMPS program office at Emory University. We thank KEMRI-Wellcome Trust Research Programme, Kilifi, Kenya, and the hospital administration and staff at both Hiwot Fana Comprehensive Specialised Hospital (HFCSH), and Kilifi County Hospital (KCH). We are grateful to study participants and their families.

## Disclaimer

The findings and conclusions in this report are those of the author(s) and do not necessarily represent the views of the U.S. Centers for Disease Control and Prevention.

