## Supplementary material for "Effect of maternal infection on stillbirths and early neonatal deaths: nested case-control studies in pregnancy cohorts in East Africa"

Supplementary Table 12: Prevalence of bacteraemia (conventional blood culture) by species in maternal participants and odds ratio for perinatal death, Hiwot Fana Comprehensive Specialised Hospital, Ethiopia. ..16

Supplementary Table 1: Summary of Maternal Infection study design, by site, data and sample collection and laboratory testing.

| **Site** | **Kilifi County Hospital, Kenya** | **Hiwot Fana Comprehensive Specialized University Hospital, Harar, Ethiopia** |
|---|---|---|
| **Study design** | Retrospective observational cohort within the maternal cohort recruited 2011-2017 as part of the Kilifi Perinatal and Maternal Health Research Programme (KIPMAT) with nested case-control study | Prospective observational cohort with the maternal cohort recruited prospectively 2018-2020, with nested case-control of study |
| **Meta-data** | Maternal clinical admission records, linked laboratory and demographic data. | Maternal clinical admission records, linked laboratory investigations. |
| **Cases** | Mothers with a stillbirth ($\geq$28 weeks' gestation of >1000grams) or very early (<24hours of birth) neonatal death | |
| **Controls** | Mothers with a livebirth, surviving to >24hours selected by simple randomisation from those eligible. | |
| **Sample size** | cases=350; controls=500 | cases = 350; controls = 350 |
| **Samples** | Maternal blood (stored samples) taken at delivery. | Maternal blood taken at delivery, NP-OP swabs, VR swabs. |
| **Laboratory tests*** | Nucleic acid extraction and multiplex PCR (Taqman Array Cards). | Conventional blood culture (automated, with isolate storage at -80ºC), conventional culture of VR swab.<br>Nucleic acid extraction and multiplex PCR (Taqman Array Cards) on blood, NP-OP swabs and VR swabs. |

**NP-OP=Nasopharyngeal or oropharyngeal, VR=vagino-rectal, PCR=polymerase chain reaction, TAC=Taqman Array Card*

Supplementary Table 2: STROBE checklist

| | | Reporting Item | Page Number |
|---|---|---|---|
| **Title and abstract** | | | |
| Title | #1a | Indicate the study's design with a commonly used term in the title or the abstract | Title page |
| Abstract | #1b | Provide in the abstract an informative and balanced summary of what was done and what was found | 1 |
| **Introduction** | | | |
| Background / rationale | #2 | Explain the scientific background and rationale for the investigation being reported | 2 |
| Objectives | #3 | State specific objectives, including any prespecified hypotheses | 2 |
| **Methods** | | | |
| Study design | #4 | Present key elements of study design early in the paper | 2-4 |
| Setting | #5 | Describe the setting, locations, and relevant dates, including periods of recruitment, exposure, follow-up, and data collection | 2-3 |
| Eligibility criteria | #6a | Give the eligibility criteria, and the sources and methods of case ascertainment and control selection. Give the rationale for the choice of cases and controls. For matched studies, give matching criteria and the number of controls per case | 3-4 |
| Eligibility criteria | #6b | For matched studies, give matching criteria and the number of controls per case | N/A |
| | #7 | Clearly define all outcomes, exposures, predictors, potential confounders, and effect modifiers. Give diagnostic criteria, if applicable | 3-4 |
| Data sources / measurement | #8 | For each variable of interest give sources of data and details of methods of assessment (measurement). Describe comparability of assessment methods if there is more than one group. Give information separately for cases and controls. | 3-4 |
| Bias | #9 | Describe any efforts to address potential sources of bias | 5 |
| Study size | #10 | Explain how the study size was arrived at | 4 |
| Quantitative variables | #11 | Explain how quantitative variables were handled in the analyses. If applicable, describe which groupings were chosen, and why | 4 |
| Statistical methods | #12a | Describe all statistical methods, including those used to control for confounding | 4 |
| Statistical methods | #12b | Describe any methods used to examine subgroups and interactions | 4 |
| Statistical methods | #12c | Explain how missing data were addressed | 4 |
| Statistical methods | #12d | If applicable, explain how matching of cases and controls was addressed | N/A |
| Statistical methods | #12e | Describe any sensitivity analyses | 4 |
| **Results** | | | |
| Participants | #13a | Report numbers of individuals at each stage of study—eg numbers potentially eligible, examined for eligibility, confirmed eligible, included in the study, completing follow-up, and analysed. Give information separately for cases and controls. | 6, Fig 1 |
| Participants | #13b | Give reasons for non-participation at each stage | 6, Fig 1 |
| Participants | #13c | Consider use of a flow diagram | Fig 1 |
| Descriptive data | #14a | Give characteristics of study participants (eg demographic, clinical, social) and information on exposures and potential confounders. Give information separately for cases and controls | Table 1 |
| Descriptive data | #14b | Indicate number of participants with missing data for each variable of interest | Table 1, Supplementary tables |

| | | | |
|---|---|---|---|
| Outcome data | #15 | Report numbers in each exposure category, or summary measures of exposure. Give information separately for cases and controls | Table 1, Supplementary tables |
| Main results | #16a | Give unadjusted estimates and, if applicable, confounder-adjusted estimates and their precision (eg, 95% confidence interval). Make clear which confounders were adjusted for and why they were included | Table 1, Supplementary tables. P5. |
| Main results | #16b | Report category boundaries when continuous variables were categorized | Table 1 |
| Main results | #16c | If relevant, consider translating estimates of relative risk into absolute risk for a meaningful time period | N/A |
| Other analyses | #17 | Report other analyses done—e.g., analyses of subgroups and interactions, and sensitivity analyses | Supplementary tables |
| **Discussion** | | | |
| Key results | #18 | Summarise key results with reference to study objectives | 8 |
| Limitations | #19 | Discuss limitations of the study, taking into account sources of potential bias or imprecision. Discuss both direction and magnitude of any potential bias. | 8-9 |
| Interpretation | #20 | Give a cautious overall interpretation considering objectives, limitations, multiplicity of analyses, results from similar studies, and other relevant evidence. | 8-9 |
| Generalisability | #21 | Discuss the generalisability (external validity) of the study results | 9 |
| **Other Information** | | | |
| Funding | #22 | Give the source of funding and the role of the funders for the present study and, if applicable, for the original study on which the present article is based | 5 |

Supplementary Table 3: Infectious targets tested for using TaqMan Array Cards for each maternal sample types (blood, vagino-rectal swab, oropharyngeal swab). Grey shading indicates that the target was included in the card for that specimen type **

| **Bacteria** | **Blood** | **VRS*** | **OPS*** | **Viruses** | **Blood** | **VRS*** | **OPS*** |
|---|---|---|---|---|---|---|---|
| *Acinetobacter baumanii* | | | | Adenovirus | | | |
| *Bartonella spp.* | | | | Crimean-Congo hemorrhagic fever virus | | | |
| *Bordatella parapertussis* | | | | Chickungunya virus | | | |
| *Bordatella pertussis* | | | | hCoV 229E | | | |
| *Brucella spp.* | | | | hCoV HKU1 | | | |
| *Burkholderia pseudommallei* | | | | hCoV NL63 | | | |
| *Corynebacterium. diphtheriae* | | | | hCoV OC43 | | | |
| *Corynebacterium pneumoniae* | | | | Cytomegalovirus | | | |
| *Corynebacterium ulcerans/pseudotb* | | | | Dengue virus | | | |
| *Coxiella burnetii* | | | | Enterovirus | | | |
| *Chlamydia trachomatis* | | | | Hepatitis E virus | | | |
| DT (toxin) | | | | Human metapneumovirus | | | |
| *Escherichia coli/Shigella spp.* | | | | Herpes simplex virus 1 | | | |
| *Enterococcus faecium* | | | | Herpes simplex virus 2 | | | |
| *Enterococcus faecalis* | | | | Influenza A | | | |
| *group A streptococcus* | | | | Influenza B | | | |
| *group B streptococcus* | | | | Japanese encephalitis virus | | | |
| *Haemophilus influenzae* type b | | | | Lassa A | | | |
| *Haemophilus influenzae* | | | | Lassa B | | | |
| *Klebsiella pneumoniae* | | | | Lassa virus (pan) | | | |
| *Listeria monocytogenes* | | | | Measles virus | | | |
| *Leptospira spp* | | | | MERS Coronavirus | | | |
| *Moraxella catarrhalis* | | | | MERS-CoV upE | | | |
| *Mycobacterium tuberculosis* | | | | Mumps virus | | | |
| *Mycoplasma pneumoniae* | | | | Nipah virus | | | |
| *Neisseria meningitidis* | | | | Parechovirus | | | |
| *Neisseria gonorrhoeae* | | | | Parainfluenza 1 | | | |
| *Orientia tsutsugamushi* | | | | Parainfluenza 2 | | | |
| *Pseudomonas aeruginosa* | | | | Parainfluenza 3 | | | |
| *Salmonella enterica* serovar Paratyphi A | | | | Parainfluenza 4 | | | |
| *Streptococcus pneumoniae* | | | | Parvovirus B19 virus | | | |
| *Streptococcus suis* | | | | Rubella virus | | | |
| *Salmonella spp.* | | | | Respiratory syncytial virus | | | |
| *S. enterica* serovar Typhi | | | | Rhinovirus | | | |
| *Staphylococcus aureus* | | | | Rift valley fever virus | | | |
| *Treponema pallidum* | | | | Varicella zoster virus | | | |
| *Ureaplasma spp.* | | | | West nile virus | | | |
| *Yersinia spp.* | | | | Yellow fever virus | | | |
| *Rickettsia spp.* | | | | Zika virus | | | |
| **Fungi** | | | | **Protozoa** | | | |
| *Candida albicans* | | | | *Plasmodium falciparum* | | | |
| *Cryptococcus neoformans/gattii* | | | | *Plasmodium vivax* | | | |
| *Pneumocystis jirovecii* | | | | *Toxoplasma gondii* | | | |

*VRS=vagino rectal swab; OPS= oropharyngeal swab

**Maureen H Diaz, Jessica L Waller et al. Development and Implementation of Multiplex TaqMan Array Cards for Specimen Testing at Child Health and Mortality Prevention Surveillance Site Laboratories, *Clinical Infectious Diseases*, Volume 69, Issue Supplement_4, 15 October 2019, Pages S311 - S321, https://doi.org/10.1093/cid/ciz571

Supplementary Table 4: Total cases required to detect different effect sizes (odds ratios) against the prevalence of maternal infection in controls

| | Prevalence of exposure in controls | | | | | | | | |
|---|---|---|---|---|---|---|---|---|---|
| Odds ratio | **0.001** | **0.002** | **0.005** | **0.01** | **0.02** | **0.04** | **0.06** | **0.08** | **0.1** |
| **2** | 20912 | 10476 | 4214 | 2127 | 1084 | 563 | 389 | 304 | 252 |
| **3** | 6920 | 3469 | 1398 | 708 | 363 | 191 | 134 | 106 | 89 |
| **4** | 3828 | 1920 | 776 | 394 | 203 | 108 | 76 | 61 | 52 |
| 6 | 1921 | 964 | 391 | 200 | 104 | 56 | 41 | 33 | 28 |
| **8** | 1257 | 632 | 257 | 132 | 69 | 38 | 28 | 23 | 20 |

| | |
|---|---|
| (dark grey) | Over 700 cases, study not powered to detect these odds ratios with this prevalence of maternal infection in controls |
| (light grey) | Over 350 cases, study not powered to detect these odds ratios with this prevalence of maternal infection in controls |
| | Assume 4:5 ratio (cases:controls), alpha=0.05, power=80% |

Supplementary Table 5: Maternal and newborn characteristics in cases and controls associated with perinatal mortality, Kilifi County Hospital, Kenya

| | Univariable analyses | | | Multivariable analyses | | |
|---|---|---|---|---|---|---|
| | **OR** | **95%CI** | **p** | **OR** | **95%CI** | **p** |
| **Age** | | | | | | |
| <20 | 0.79 | (0.52-1.19) | | 0.8 | (0.52-1.23) | |
| 20 to <30 | 1 | | | 1 | | |
| 30 to <40 | 1.39 | (1.00-1.94) | | 1.46 | (1.03-2.07) | |
| ≥40years | 2.64 | (1.16-6.03) | 0.007 | 2.49 | (1.07-5.78) | 0.011 |
| **Marital status** | | | | | | |
| Married | 1 | | | | | |
| Single | 0.99 | (0.60-1.65) | | | | |
| Divorced | 2.47 | (0.41-14.84) | | | | |
| Widowed | * | | 0.6 | | | |
| **Education level** | | | | | | |
| None | 1.45 | (0.99-2.12) | | 1.29 | (0.86-1.94) | |
| Primary | 1 | | | 1 | | |
| Secondary | 1.03 | (0.70-1.52) | | 1.04 | (0.70-1.55) | |
| Higher | 0.35 | (0.15-0.81) | 0.006 | 0.36 | (0.12-0.74) | 0.010 |
| **Nulliparous** | | | | | | |
| No | 1 | | | | | |
| Yes | 0.9 | (0.67-1.22) | 0.5 | | | |
| **History of fever** | | | | | | |
| No | 1 | | | | | |
| Yes | 2.61 | (0.85-8.06) | 0.095 | | | |
| **PROM >18h** | | | | | | |
| No | 1 | | | | | |
| Yes | 1.02 | (0.53-1.98) | 1 | | | |
| **Dysuria** | | | | | | |
| No | 1 | | | | | |
| Yes | 0.32 | (0.09-1.10) | 0.07 | | | |
| **Positive nitrite and/or leucocytes** | | | | | | |
| No | 1 | | | | | |
| Yes | 0.83 | (0.58-1.18) | 0.3 | | | |
| **Emergency clinical signs** | | | | | | |
| No | 1 | | | | | |
| Yes | 2.98 | (2.06-4.31) | <0.001 | 2.89 | (1.98-4.24) | <0.001 |
| **Mode of delivery** | | | | | | |
| Vaginal | 1 | | | | | |
| Caesarean section | 1 | (0.70-1.43) | 1 | | | |
| **Birthweight (g)** | | | | | | |
| 1000-1500 | 19.62 | (9.14-42.13) | | 18.63 | (8.65-40.14) | |
| 1500- 2500 | 3.04 | (2.13-4.34) | | 2.93 | (2.04-4.21) | |
| Over 2500 | 1 | | <0.001 | 1 | | <0.001 |
| **Sex** | | | | | | |
| Male | 1.24 | (0.93-1.66) | 0.1 | 1.13 | (0.82-1.55) | 0.5 |
| Female | 1 | | | 1 | | |

*insufficient data

Supplementary Table 6: Maternal and newborn characteristics in cases and controls associated with perinatal mortality, Hiwot Fana Comprehensive Specialized Hospital, Ethiopia

| | **Univariable analyses** | | | **Multivariable analyses** | | |
|---|---|---|---|---|---|---|
| | **OR** | **95%CI** | **p** | **OR** | **95%CI** | **p** |
| **Age** | | | | | | |
| <20 | 1.45 | (0.95-2.50) | | 1.19 | (0.66-2.17) | |
| 20 to <30 | 1 | | | 1 | | |
| 30 to <40 | 1.88 | (1.30-2.73) | | 1.37 | (0.91-2.07) | |
| ≥40years | 0.66 | (0.22-1.97) | 0.004 | 0.35 | (0.11-1.07) | 0.072 |
| **Marital status** | | | | | | |
| Married | 1 | | | | | |
| Single | 1.05 | (0.07-16.90) | | | | |
| Divorced | * | | | | | |
| Widowed | * | | 1 | | | |
| **Education level** | | | | | | |
| None | 3.88 | (2.51-6.00) | | 3.81 | (2.40-6.03) | |
| Primary | 1 | (0.38-1.48) | | | | |
| Secondary | 0.75 | (0.38-1.48) | | 0.76 | (0.38-1.53) | |
| Higher | 1.6 | (0.84-3.03) | <0.001 | 1.69 | (0.86-3.32) | <0.001 |
| **Nulliparous** | | | | | | |
| No | 1 | | | | | |
| Yes | 0.83 | (0.18-3.73) | 0.8 | | | |
| **History of fever** | | | | | | |
| No | 1 | | | | | |
| Yes | 1.73 | (0.71-4.22) | 0.8 | | | |
| **PROM >18h** | | | | | | |
| No | 1 | | | | | |
| Yes | 0.91 | (0.57-1.46) | 0.7 | | | |
| **Dysuria** | | | | | | |
| No | 1 | | | | | |
| Yes | 0.55 | (0.05-6.15) | 0.6 | | | |
| **Positive nitrite and/or leucocytes** | | | | | | |
| No | 1 | | | | | |
| Yes | 0.96 | (0.59-1.55) | 0.9 | | | |
| **Emergency clinical signs** | | | | | | |
| No | 1 | | | 1 | | |
| Yes | 1.84 | (1.35-2.52) | <0.001 | 1.53 | (1.07-2.20) | 0.021 |
| **Mode of delivery** | | | | | | |
| Vaginal | 1 | | | | | |
| Caesarean section | 0.85 | (0.59-1.20) | 0.4 | | | |
| **Birthweight (g)** | | | | | | |
| 1000-1500 | 17.39 | (7.30-41.46) | | 17.67 | (7.38-42.31) | |
| 1500- 2500 | 5.73 | (3.57-9.22) | | 5.75 | (3.53-9.37) | |
| Over 2500 | 1 | | <0.001 | 1 | | <0.001 |
| **Sex** | | | | | | |
| Male | 1.18 | (0.85-1.62) | 0.3 | 1.37 | (0.94-1.98) | 0.098 |
| Female | 1 | | | 1 | | |

*insufficient data

Supplementary Table 7: Maternal and newborn characteristics in cases and controls associated with perinatal mortality, combined analysis for Kilifi County Hospital, Kenya and Hiwot Fana Comprehensive Specialized Hospital, Ethiopia

| | Univariable analyses | | | Multivariable analyses | | |
|---|---|---|---|---|---|---|
| | **OR** | **95%CI** | **p** | **OR** | **95%CI** | **p** |
| **Age** | | | | | | |
| <20 | 0.98 | (0.71-1.35) | | 0.92 | (0.65-1.30) | |
| 20 to <30 | 1 | | | 1 | | |
| 30 to <40 | 1.59 | (1.25-2.03) | | 1.42 | (1.09-1.84) | |
| ≥40years | 1.60 | (0.84-3.02) | 0.001 | 1.11 | (0.56-2.20) | 0.046 |
| **Marital status** | | | | | | |
| Married | 1 | | | | | |
| Single | 0.97 | (0.59-1.61) | | | | |
| Divorced | 2.41 | (0.40-14.51) | | | | |
| Widowed | | | 0.6 | | | |
| **Education level** | | | | | | |
| None | 2.42 | (1.90-3.09) | | 1.90 | (1.46-2.48) | |
| Primary | 1 | | | 1 | | |
| Secondary | 0.91 | (0.65-1.27) | | 0.85 | (0.60-1.19) | |
| Higher | 0.80 | (0.51-1.26) | <0.001 | 0.63 | (0.38-1.04) | <0.001 |
| **Nulliparous** | | | | | | |
| No | 1 | | | | | |
| Yes | 0.92 | (0.74-1.16) | 0.5 | | | |
| **History of fever** | | | | | | |
| No | 1 | | | | | |
| Yes | 1.73 | (0.71-4.22) | 0.2 | | | |
| **PROM >18h** | | | | | | |
| No | 1 | | | | | |
| Yes | 0.96 | (0.66-1.41) | 0.8 | | | |
| **Dysuria** | | | | | | |
| No | 1 | | | 1 | | |
| Yes | 0.35 | (0.12-1.03) | 0.058 | 0.38 | (0.12-1.18) | 0.095 |
| **Positive nitrite and/or leucocytes** | | | | | | |
| No | 1 | | | | | |
| Yes | 0.88 | (0.66-1.17) | 0.4 | | | |
| **Emergency clinical signs** | | | | | | |
| No | 1 | | | 1 | | |
| Yes | 2.33 | (1.88-2.89) | <0.001 | 1.91 | (1.51-2.43) | <0.001 |
| **Mode of delivery** | | | | | | |
| Vaginal | 1 | | | | | |
| Caesarean section | 0.92 | (0.72-1.19) | 0.5 | | | |
| **Birthweight (g)** | | | | | | |
| 1000-1500 | 18.8 | (10.58-33.41) | | 18.3 | (10.24-32.51) | |
| 1500- 2500 | 3.84 | (2.90-5.08) | | 3.72 | (2.80-4.94) | |
| Over 2500 | 1 | | <0.001 | 1 | | <0.001 |
| **Sex of the baby** | | | | | | |
| Male | 1.22 | (0.98-1.51) | 0.071 | 1.21 | (0.97-1.54) | 0.1 |
| Female | 1 | | | 1 | | |

Supplementary Table 8: Maternal and newborn characteristics in cases and controls associated with maternal bacteraemia (conventional blood culture) in Hiwot Fana Comprehensive Specialized Hospital, Ethiopia

| | Univariable analyses | | | Multivariable analyses | | |
|---|---|---|---|---|---|---|
| | **OR** | **95%CI** | **p** | **OR** | **95%CI** | **p** |
| **Age** | | | | | | |
| <20 | 0.96 | (0.28-3.34) | | 0.91 | (0.25-3.22) | |
| 20 to <30 | 1 | | | 1 | | |
| 30 to <40 | 1.24 | (0.56-2.72) | | 1.15 | (0.50-2.64) | |
| ≥40years | 1.37 | (0.17-11.0) | 0.9 | 1.26 | (0.15-10.31) | 1 |
| **Marital status** | | | | | | |
| Married | 1 | | | | | |
| Single | * | | | | | |
| Divorced | * | | | | | |
| Widowed | * | | | | | |
| **Education level** | | | | | | |
| None | 1.42 | (0.57-3.58) | | | | |
| Primary | 1 | | | | | |
| Secondary | 0.96 | (0.23-3.97) | | | | |
| Higher | 0.71 | (0.14-3.61) | 0.7 | | | |
| **Nulliparous** | | | | | | |
| No | 1 | | | | | |
| Yes | 1.52 | (0.72-3.22) | 0.3 | | | |
| **History of fever** | | | | | | |
| No | 1 | | | 1 | | |
| Yes | 7.73 | (1.44-41.5) | 0.017 | 6.12 | (1.11-33.00) | 0.037 |
| **PROM >18h** | | | | | | |
| No | 1 | | | | | |
| Yes | 0.73 | (0.22-2.47) | 0.6 | | | |
| **Dysuria** | | | | | | |
| No | 1 | | | | | |
| Yes | * | | | | | |
| **Positive nitrite and/or leucocytes** | | | | | | |
| No | 1 | | | | | |
| Yes | 1.24 | (0.36-4.21) | 0.7 | | | |
| **Emergency clinical signs** | | | | | | |
| No | 1 | | | 1 | | |
| Yes | 2.44 | (1.09-5..47) | 0.031 | 2.13 | (1.11-5.08) | 0.087 |
| **Mode of delivery** | | | | | | |
| Vaginal | 1 | | | | | |
| Caesarean section | 1.31 | (0.61-2.79) | 0.49 | | | |
| **Birthweight (g)** | | | | | | |
| 1000-1500 | 1.25 | (0.42-3.75) | | | | |
| 1500- 2500 | 0.48 | (0.14-1.64) | | | | |
| Over 2500 | 1 | | 0.4 | | | |
| **Sex** | | | | | | |
| Male | 1.06 | (0.50-2.22) | 0.9 | | | |
| Female | 1 | | | | | |

*insufficient data

Supplementary Table 9: Odds ratios for association of bacteraemia (clinically significant blood culture) with perinatal death in Hiwot Fana Comprehensive Specialised Hospital; univariable and multivariable analyses.

| | **Univariable analyses** | | | **Multivariable analyses** | | |
|---|---|---|---|---|---|---|
| | **OR** | **95%CI** | **p** | **OR** | **95%CI** | **p** |
| **Bacteraemia** | | | | | | |
| No | 1 | | | 1 | | |
| Yes | **2.70** | **(1.27-5.73)** | **0.01** | **3.67** | **(1.46-9.23)** | **0.006** |
| **Age** | | | | | | |
| <20 | 1.45 | (0.95-2.50) | 0.004 | 1.29 | (0.70-2.36) | 0.045 |
| 20 to <30 | 1 | | | 1 | | |
| 30 to <40 | 1.88 | (1.30-2.73) | | 1.54 | (1.00-2.35) | |
| ≥40years | 0.66 | (0.22-1.97) | | 0.39 | (0.12-1.23) | |
| **Marital status** | | | | | | |
| Married | 1 | | 1 | | | |
| Single | 1.05 | (0.07-16.90) | | | | |
| Divorced | * | | | | | |
| Widowed | * | | | | | |
| **Education level** | | | | | | |
| None | 3.88 | (2.51-6.00) | <0.001 | 3.77 | (2.3206.12) | <0.001 |
| Primary | 1 | (0.38-1.48) | | 1 | | |
| Secondary | 0.75 | (0.38-1.48) | | 0.65 | (0.31-1.38) | |
| Higher | 1.6 | (0.84-3.03) | | 1.74 | (0.85-1.53) | |
| **Nulliparous** | | | | | | |
| No | 1 | | | | | |
| Yes | 0.96 | (0.68-1.35) | 0.8 | | | |
| **Sex fetus/baby** | | | | | | |
| Male | 1.18 | (0.85-1.62) | 0.3 | 1.01 | (0.70-1.45) | 1 |
| Female | 1 | | | 1 | | |

*insufficient data

Supplementary Table 10: Odds ratios for association of bacterial detection (PCR) with perinatal death in Kilifi County Hospital, in a subset of controls with "good" pregnancy outcomes; univariable and multivariable analyses.

| | Univariable analyses | | | Multivariable analyses | | |
|---|---|---|---|---|---|---|
| | **OR** | **95%CI** | **p** | **OR** | **95%CI** | **p** |
| **Bacteria in blood on PCR** | | | | | | |
| No | 1 | | | 1 | | |
| **Yes** | **2.49** | **(1.17-5.30)** | **0.018** | 2.69 | (1.21-6.02) | **0.016** |
| **Age** | | | | | | |
| <20 | 0.80 | (0.51-1.25) | | 0.72 | (0.44-1.18) | |
| 20 to <30 | 1 | | | 1 | | |
| 30 to <40 | 1.45 | (1.00-2.10) | | 1.51 | (1.02-2.24) | |
| ≥40years | 1.64 | (0.72-3.76) | 0.056 | 1.28 | (0.54-3.08) | 0.042 |
| **Marital status** | | | | | | |
| Married | 1 | | | | | |
| Single | 0.85 | (0.49-1.46) | | | | |
| Divorced | 3.03 | (0.31-29.3) | | | | |
| Widowed | * | | 0.5 | | | |
| **Education level** | | | | | | |
| None | 1.39 | (0.91-2.13) | | 1.25 | (0.80-1.97) | |
| Primary | 1 | | | 1 | | |
| Secondary | 1.13 | (0.73-1.73) | | 1.15 | (0.73-1.82) | |
| Higher | 0.38 | (0.15-0.94) | 0.039 | 0.36 | (0.14-0.95) | 0.077 |
| **Nulliparous** | | | | | | |
| No | 1 | | | | | |
| Yes | 1.04 | (0.74-1.44) | 0.8 | | | |
| **Sex fetus/baby** | | | | | | |
| Male | 1.34 | (0.97-1.85) | 0.073 | 1.30 | (0.93-1.82) | 0.1 |
| Female | 1 | | | 1 | | |

*insufficient data

Supplementary Table 11: Odds ratios for association of *Escherichia coli* (conventional blood culture or PCR) with perinatal death in Hiwot Fana Comprehensive Specialised Hospital and Kilifi County Hospital, univariable and multivariable analyses

| | **Univariable analyses** | | | **Multivariable analyses** | | |
|---|---|---|---|---|---|---|
| | **OR** | **95%CI** | **p** | **OR** | **95%CI** | **p** |
| ***E Coli* PCR/culture** | | | | | | |
| No | 1 | | | 1 | | |
| Yes | **2.19** | **(1.00-4.81)** | **0.05** | **2.62** | **(1.08-6.32)** | **0.032** |
| **Age** | | | | | | |
| <20 | 0.98 | (0.71-1.35) | | 0.90 | (0.64-1.27) | |
| 20 to <30 | 1 | | | 1 | | |
| 30 to <40 | 1.59 | (1.25-2.03) | | 1.44 | (1.11-1.88) | |
| ≥40years | 1.60 | (0.84-3.02) | 0.001 | 1.09 | (0.55-2.15) | 0.028 |
| **Marital status** | | | | | | |
| Married | 1 | | | | | |
| Single | 0.97 | (0.59-1.61) | | | | |
| Divorced | 2.41 | (0.40-14.51) | | | | |
| Widowed | | | 0.6 | | | |
| **Education level** | | | | | | |
| None | 2.42 | (1.90-3.09) | | 2.26 | (1.74-2.93) | |
| Primary | 1 | | | | | |
| Secondary | 0.91 | (0.65-1.27) | | 0.88 | (0.62-1.24) | |
| Higher | 0.80 | (0.51-1.26) | <0.001 | 0.75 | (0.46-1.23) | <0.001 |
| **Nulliparous** | | | | | | |
| No | 1 | | | | | |
| Yes | 0.92 | (0.74-1.16) | 0.5 | | | |
| **Sex** | | | | | | |
| Male | 1.22 | (0.98-1.51) | 0.071 | 1.14 | (0.91-1.44) | 0.3 |
| Female | 1 | | | 1 | | |

Supplementary Table 12: Prevalence of bacteraemia (conventional blood culture) by species in maternal participants and odds ratio for perinatal death, Hiwot Fana Comprehensive Specialised Hospital, Ethiopia.

| | **Case** | | **Control** | | **Fisher's exact test** | **Odds Ratio** | **(95% CI)** | **p** |
|---|---|---|---|---|---|---|---|---|
| | **n=324** | **(%)** | **n=347** | **(%)** | | | | |
| *Aeromonas hydrophila* | 4 | 1.2 | 2 | 0.6 | 0.4 | 2.2 | (0.4-11.9) | 0.4 |
| *Brevimundus vesicularis* | 0 | 0.0 | 2 | 0.6 | 0.5 | * | | |
| *Burkholderia cepacia* | 3 | 0.9 | 0 | 0.0 | 0.1 | ** | | |
| *Cupriavidus pauculus* | 0 | 0.0 | 1 | 0.3 | 1.0 | * | | |
| *Escherichia coli/Shigella spp.* | 5 | 1.5 | 0 | 0.0 | 0.026 | ** | | |
| group A streptococcus | 1 | 0.3 | 0 | 0.0 | 0.5 | ** | | |
| group B streptococcus | 2 | 0.6 | 0 | 0.0 | 0.2 | ** | | |
| group C/G streptococcus | 3 | 0.9 | 0 | 0.0 | 0.1 | ** | | |
| group D streptococcus | 3 | 0.9 | 1 | 0.3 | 0.4 | 3.2 | (0.3-31.3) | 0.3 |
| *Streptococcus spp.* | 0 | 0.0 | 1 | 0.3 | 1.0 | * | | |
| *Neisseria gonorrhoeae* | 2 | 0.6 | 1 | 0.3 | 0.6 | 2.1 | (0.2-23.8) | 0.5 |
| *Salmonella enterica* serovar Paratyphi A | 1 | 0.3 | 0 | 0.0 | 0.5 | ** | | |
| *Salmonella spp.* | 1 | 0.3 | 0 | 0.0 | 0.5 | ** | | |
| *Staphylococcus aureus* | 4 | 1.2 | 2 | 0.6 | 0.4 | 2.2 | (0.4-11.9) | 0.4 |

* bacteraemia detected only in controls ** bacteraemia detected only in cases

Supplementary Table 13: Prevalence of each bacterial, viral, fungal, parasitic target molecularly detected (PCR) in blood in maternal participants, by species, and odds ratio for perinatal death, Kilifi County Hospital, Kenya.

| | Case | | Control | | Fisher's exact test | Odds Ratio | (95% CI) | p |
|---|---|---|---|---|---|---|---|---|
| | N=318 | | N=508 | | | | | |
| **Bacteria** | | | | | | | | |
| *Acinetobacter baumanii* | 0 | 0.0 | 1 | 0.2 | 0.6 | * | | |
| *Bartonella spp.* | 0 | 0.0 | 0 | 0 | | | | |
| *Brucella spp.* | 5 | 1.6 | 5 | 1 | 0.5 | 1.6 | (0.5-5.6) | 0.5 |
| *Burkholderia pseudommallei* | 0 | 0.0 | 0 | 0 | | | | |
| *Coxiella burnetii* | 1 | 0.3 | 2 | 0.4 | 1 | 0.8 | (0.1-8.8) | 0.9 |
| *Escherichia coli/Shigella spp.* | 4 | 1.3 | 2 | 0.4 | 0.2 | 3.2 | (0.6-17.7) | 0.2 |
| *Enterococcus faecium* | 1 | 0.3 | 1 | 0.2 | 1 | 1.6 | (0.1-25.7) | 0.7 |
| *Enterococcus faecalis* | 2 | 0 | 2 | 0.4 | 0.6 | 1.6 | (0.2-11.4) | 0.6 |
| group A streptococcus | 0 | 0.0 | 0 | 0 | | | | |
| group B streptococcus | 0 | 0.0 | 0 | 0 | | | | |
| *H. influenzae* type b | 0 | 0.0 | 0 | 0 | | | | |
| *Haemophilus influenzae* | 0 | 0.0 | 0 | 0 | | | | |
| *Klebsiella pneumoniae* | 0 | 0.0 | 0 | 0 | | | | |
| *Listeria monocytogenes* | 0 | 0.0 | 0 | 0 | | | | |
| *Leptospira spp* | 0 | 0.0 | 0 | 0 | | | | |
| *Moraxella catarrhalis* | 0 | 0.0 | 5 | 1 | 0.2 | * | | |
| *Mycobacterium tuberculosis* | 1 | 0.3 | 2 | 0.4 | 1 | 0.8 | (0.1-2.8) | 0.9 |
| *Neisseria meningitidis* | 0 | 0.0 | 0 | 0 | | | | |
| *Neisseria gonorrhoeae* | 0 | 0.0 | 1 | 0.2 | 1 | * | | |
| *Orientia tsutsugamushi* | 0 | 0.0 | 0 | 0 | | | | |
| *Pseudomonas aeruginosa* | 2 | 0.6 | 4 | 0.8 | 1 | 0.8 | (0.2-4.4) | 0.8 |
| *Streptococcus pneumoniae* | 1 | 0.3 | 3 | 0.6 | 1 | 0.5 | (0.1-5.1) | 0.6 |
| *Streptococcus suis* | 0 | 0.0 | 0 | 0 | | | | |
| *Salmonella enterica* serovar Paratyphi A | 1 | 0.3 | 0 | 0 | 0.4 | ** | | |
| Salmonella spp. | 1 | 0.3 | 0 | 0 | 0.4 | ** | | |
| *Salmonella enterica* serovar Typhi | 0 | 0.0 | 0 | 0 | | | | |
| *Staphylococcus aureus* | 1 | 0.3 | 0 | 0 | 0.4 | ** | | |
| *Treponema pallidum* | 1 | 0.3 | 3 | 0.6 | 1 | 0.5 | (0.1-5.1) | 0.6 |
| *Ureaplasma spp.* | 0 | 0.0 | 2 | 0.4 | 0.5 | * | | |
| *Yersinia spp.* | 0 | 0 | 0 | 0 | | | | |
| *Rickettsia spp.* | 3 | 0.9 | 1 | 0.2 | 0.2 | 4.8 | (0.5-46.2) | 0.2 |
| **Viruses** | | | | | | | | |
| Adenovirus | 3 | 0.9 | 2 | 0.4 | 0.4 | 2.4 | (0.4-14.5) | 0.3 |
| Crimean-Congo hemorrhagic fever virus | 0 | 0.0 | 0 | 0 | | | | |
| Chikungunya virus | 3 | 0.9 | 1 | 0.2 | 0.2 | 4.8 | (0.5-46.6) | 0.2 |
| Cytomegalovirus | 8 | 2.5 | 9 | 1.8 | 0.5 | 1.4 | (0.6-3.8) | 0.5 |
| Dengue virus | 2 | 0.6 | 2 | 0.4 | 0.6 | 1.6 | (0.2-11.4) | 0.6 |
| Enterovirus | 0 | 0.0 | 0 | 0 | | | | |
| Hepatitis E virus | 0 | 0.0 | 0 | 0 | | | | |

| | | | | | | | | |
|---|---|---|---|---|---|---|---|---|
| Herpes simplex virus 1 | 0 | 0.0 | 0 | 0 | | | | |
| Herpes simplex virus 2 | 0 | 0.0 | 0 | 0 | | | | |
| Japanese encephalitis virus | 0 | 0.0 | 0 | 0 | | | | |
| Lassa A | 0 | 0.0 | 0 | 0 | | | | |
| Lassa B | 0 | 0.0 | 0 | 0 | | | | |
| Lassa virus (pan) | 2 | 0.6 | 3 | 0.6 | 1 | 1.07 | (0.2-6.4) | 0.9 |
| Measles virus | 0 | 0.0 | 1 | 0.2 | 1 | * | | |
| Mumps virus | 0 | 0.0 | 0 | 0 | | | | |
| Nipah virus | 0 | 0.0 | 1 | 0.2 | 1 | * | | |
| Parechovirus | 0 | 0.0 | 0 | 0 | | | | |
| Parvovirus B19 | 4 | 1.3 | 4 | 0.8 | 0.5 | 1.6 | (0.4-6.5) | 0.5 |
| Rubella virus | 8 | 2.5 | 19 | 3.7 | 0.4 | 0.7 | (0.3-1.5) | 0.3 |
| Rift valley fever virus | 0 | 0.0 | 0 | 0 | | | | |
| Varicella zoster virus | 1 | 0.3 | 0 | 0 | 0.4 | ** | | |
| West Nile virus | 0 | 0.0 | 0 | 0 | | | | |
| Yellow fever virus | 0 | 0.0 | 0 | 0 | | | | |
| Zika virus | 2 | 0.6 | 2 | 0.4 | 0.6 | 1.6 | (0.2-11.4) | 0.6 |
| **Protozoa** | | | | | | | | |
| *Plasmodium falciparum* | 13 | 4.1 | 24 | 4.7 | 0.7 | 0.9 | (0.4-1.7) | 0.7 |
| *Plasmodium vivax* | 0 | 0.0 | 0 | 0 | | | | |
| *Toxoplasma gondii* | 0 | 0.0 | 0 | 0 | | | | |
| **Fungi** | | | | | | | | |
| *Candida albicans* | 0 | 0.0 | 2 | 0.4 | 0.5 | * | | |
| *Cryptococcus neoformans/gattii* | 0 | 0.0 | 0 | 0 | | | | |

* target detected only in controls ** target detected only in cases

Supplementary Table 14: Prevalence of each bacterial, viral, fungal, parasitic species molecularly detected (PCR) in maternal blood and odds ratio for perinatal death, Hiwot Fana Comprehensive Specialised Hospital, Ethiopia.

| | **Case N=324** | | **Control N=347** | | **Fisher's exact test** | **Odds Ratio** | **95% CI** | **p** |
|---|---|---|---|---|---|---|---|---|
| | **n** | **(%)** | **n** | **(%)** | | | | |
| **Bacteria** | | | | | | | | |
| *Acinetobacter baumanii* | 0 | 0 | 0 | 0.0 | | | | |
| *Bartonella spp.* | 0 | 0 | 0 | 0.0 | | | | |
| *Brucella spp.* | 16 | 5 | 12 | 3.5 | 0.4 | 1.5 | (0.7-3.1) | 0.3 |
| *Burkholderia pseudomallei* | 0 | 0 | 0 | 0.0 | | | | |
| *Coxiella burnetii* | 2 | 1 | 0 | 0.0 | 0.2 | 0.9 | (0.8-1.1) | 0.3 |
| *E.coli/Shigella spp.* | 10 | 3 | 8 | 2.3 | 0.6 | 1.4 | (0.5-3.5) | 0.5 |
| *Enterococcus faecium* | 0 | 0 | 0 | 0.0 | | | | |
| *Enterococcus faecalis* | 3 | 1 | 4 | 1.2 | 1.0 | 0.8 | (0.2-3.6) | 0.8 |
| group A streptococcus | 1 | 0 | 0 | 0.0 | 0.5 | | | |
| group B streptococcus | 1 | 0 | 1 | 0.3 | 1.0 | 1.1 | (0.1-17.2) | 1.0 |
| *H. influenzae type B* | 0 | 0 | 0 | 0.0 | | | | |
| *Haemophilus influenzae* | 0 | 0 | 0 | 0.0 | | | | |
| *Klebsiella pneumoniae* | 0 | 0 | 0 | 0.0 | | | | |
| *Listeria monocytogenes* | 0 | 0 | 0 | 0.0 | | | | |
| *Leptospira spp* | 0 | 0 | 0 | 0.0 | | | | |
| *Moraxella catarrhalis* | 0 | 0 | 0 | 0.0 | | | | |
| *Mycobacterium tuberculosis* | 0 | 0 | 0 | 0.0 | | | | |
| *Neisseria meningitidis* | 0 | 0 | 0 | 0.0 | | | | |
| *Neisseria gonorrhoeae* | 0 | 0 | 0 | 0.0 | | | | |
| *Orientia tsutsugamushi* | 0 | 0 | 0 | 0.0 | | | | |
| *Pseudomonas aeruginosa* | 12 | 4 | 15 | 4.3 | 0.7 | 0.9 | (0.4-1.) | 0.7 |
| *Streptococcus pneumoniae* | 1 | 0 | 0 | 0.0 | 0.5 | ** | | |
| *Streptococcus suis* | 0 | 0 | 0 | 0.0 | | | | |
| *Salmonella enterica* serovar Paratyphi A | 0 | 0 | 2 | 0.6 | 0.5 | * | | |
| *Salmonella spp.* | 2 | 1 | 3 | 0.9 | 1.0 | 0.7 | (0.1-4.3) | 0.7 |
| *Salmonella enterica* serovar Typhi | 1 | 0 | 1 | 0.3 | 1.0 | 1.1 | (0.1-17.2) | 1.0 |
| *Staphylococcus aureus* | 2 | 1 | 3 | 0.9 | 1.0 | 0.7 | (0.1-4.3) | 0.7 |
| *Treponema pallidum* | 8 | 2 | 12 | 3.5 | 0.5 | 0.7 | (0.3-1.8) | 0.5 |
| *Ureaplasma spp.* | 3 | 1 | 0 | 0.0 | 0.1 | ** | | |
| *Yersinia spp.* | 0 | 0 | 0 | 0 | | | | |
| *Rickettsia spp.* | 0 | 0 | 0 | 0 | | | | |
| **Viruses** | | | | | | | | |
| Adenovirus | 7 | 2 | 14 | 4.0 | 0.2 | 0.5 | (0.2-1.3) | 0.2 |
| Crimean-Congo hemorrhagic fever virus | 0 | 0 | 0 | 0.0 | | | | |
| Chikungunya virus | 0 | 0 | 0 | 0.0 | | | | |
| Cytomegalovirus | 8 | 2 | 7 | 2.0 | 0.8 | 1.2 | (0.4-3.4) | 0.4 |
| Dengue virus | 0 | 0 | 0 | 0.0 | | | | |
| Enterovirus | 0 | 0 | 0 | 0.0 | | | | |
| Hepatitis E virus | 0 | 0 | 0 | 0.0 | | | | |
| Herpes simplex virus 1 | 0 | 0 | 0 | 0.0 | | | | |
| Herpes simplex virus 2 | 0 | 0 | 0 | 0.0 | | | | |
| Japanese encephalitis virus | 0 | 0 | 0 | 0.0 | | | | |
| Lassa A | 0 | 0 | 0 | 0.0 | | | | |
| Lassa B | 0 | 0 | 0 | 0.0 | | | | |
| Lassa virus (pan) | 0 | 0 | 0 | 0.0 | | | | |
| Measles virus | 1 | 0 | 0 | 0.0 | 0.5 | ** | | |
| Mumps virus | 0 | 0 | 0 | 0.0 | | | | |
| Nipah virus | 0 | 0 | 0 | 0.0 | | | | |

| | | | | | | | | |
|---|---|---|---|---|---|---|---|---|
| Parechovirus | 0 | 0 | 0 | 0.0 | | | | |
| Parvovirus B19 | 12 | 4 | 16 | 4.6 | 0.6 | 0.8 | (0.4-1.7) | 0.6 |
| Rubella virus | 0 | 0 | 0 | 0.0 | | | | |
| Rift Valley Fever virus | 0 | 0 | 0 | 0.0 | | | | |
| Varicella zoster virus | 0 | 0 | 1 | 0.3 | 1.0 | * | | |
| West Nile Virus | 0 | 0 | 0 | 0.0 | | | | |
| Yellow Fever Virus | 0 | 0 | 0 | 0.0 | | | | |
| Zika virus | 4 | 1 | 1 | 0.3 | 0.2 | 4.3 | (0.5-38.9) | 0.2 |
| **Protozoa** | | | | | | | | |
| *Plasmodium falciparum* | 2 | 1 | 1 | 0.3 | 0.6 | 2.1 | (0.2-23.8) | 0.6 |
| *Plasmodium vivax* | 0 | 0 | 0 | 0.0 | | | | |
| *Toxoplasma gondii* | 0 | 0 | 0 | 0.0 | | | | |
| **Fungi** | | | | | | | | |
| *Candida albicans* | 3 | 1 | 4 | 1.2 | 1.0 | 0.8 | (0.2-3.6) | 0.8 |
| *Cryptococcus neoformans/gattii* | 0 | 0 | 0 | 0.0 | | | | |

* target detected only in controls ** target detected only in cases

Supplementary Table 15: Prevalence of each bacterial, viral, fungal, parasitic target molecularly detected (PCR) in blood in maternal participants, by species, and odds ratio for perinatal death, Kilifi County Hospital, Kenya and Hiwot Fana Comprehensive Specialised Hospital, Ethiopia.

| | Case N=642 | | Control N=855 | | Fisher's exact test | Odds Ratio | (95% CI) | p |
|---|---|---|---|---|---|---|---|---|
| | n | % | n | % | | | | |
| **Bacteria** | | | | | | | | |
| *Acinetobacter baumanii* | 0 | 0.0 | 1 | 0.1 | 1.0 | * | | |
| *Bartonella spp.* | 0 | 0.0 | 0 | 0.0 | | | | |
| *Brucella spp.* | 21 | 3.3 | 17 | 2.0 | 0.1 | 1.5 | (0.-2.9) | 0.2 |
| *Burkholderia pseudommallei* | 0 | 0.0 | 0 | 0.0 | | | | |
| *Coxiella burnetii* | 3 | 0.5 | 2 | 0.2 | 0.7 | 2.0 | (0.3-12.0) | 0.4 |
| *Escherichia coli/Shigella spp.* | 14 | 2.2 | 10 | 1.2 | 0.1 | 1.7 | (0.8-3.9) | 0.2 |
| *Enterococcus faecium* | 1 | 0.2 | 1 | 0.1 | 1.0 | 1.3 | (0.1-21.3) | 0.8 |
| *Enterococcus faecalis* | 5 | 0.8 | 6 | 0.7 | 1.0 | 1.1 | (0.3-3.7) | 0.9 |
| group A streptococcus | 1 | 0.2 | 0 | 0.0 | 0.4 | | | |
| group B streptococcus | 1 | 0.2 | 1 | 0.1 | 1.0 | 1.3 | (0.1-21.2) | 0.9 |
| *Haemophilus influenzae* type b | 0 | 0.0 | 0 | 0.0 | | | | |
| *Haemophilus influenzae* | 0 | 0.0 | 0 | 0.0 | | | | |
| *Klebsiella pneumoniae* | 0 | 0.0 | 0 | 0.0 | | | | |
| *Listeria monocytogenes* | 0 | 0.0 | 0 | 0.0 | | | | |
| *Leptospira spp* | 0 | 0.0 | 0 | 0.0 | | | | |
| *Moraxella catarrhalis* | 0 | 0.0 | 5 | 0.6 | 0.075 | * | | |
| *Mycobacterium tuberculosis* | 1 | 0.2 | 2 | 0.2 | 1.0 | 0.7 | (0.1-7.8) | 0.8 |
| *Neisseria meningitidis* | 0 | 0.0 | 0 | 0.0 | | | | |
| *Neisseria gonorrhoeae* | 0 | 0.0 | 1 | 0.1 | 1.0 | * | | |
| *Orientia tsutsugamushi* | 0 | 0.0 | 0 | 0.0 | | | | |
| *Pseudomonas aeruginosa* | 14 | 2.2 | 19 | 2.2 | 1.0 | 0.9 | (0.4-1.7) | 0.7 |
| *Streptococcus pneumoniae* | 2 | 0.3 | 3 | 0.4 | 1.0 | 0.9 | (0.2-5.3) | 0.9 |
| *Streptococcus suis* | 0 | 0.0 | 0 | 0.0 | | | | |
| *Salmonella enterica* serovar Paratyphi A | 1 | 0.2 | 2 | 0.2 | 1.0 | 0.7 | (0.1-7.4) | 0.7 |
| *Salmonella spp.* | 3 | 0.5 | 3 | 0.4 | 1.0 | 1.2 | (0.3-6.3) | 0.8 |
| *Salmonella enterica* serovar Typhi | 1 | 0.2 | 1 | 0.1 | 1.0 | 1.1 | (0.1-17.2) | 1.0 |
| *Staphylococcus aureus* | 3 | 0.5 | 3 | 0.4 | 1.0 | 1.2 | (0.3-6.3) | 0.8 |
| *Treponema pallidum* | 9 | 1.4 | 15 | 1.8 | 0.7 | 0.7 | (0.3-1.6) | 0.4 |
| *Ureaplasma spp.* | 3 | 0.5 | 2 | 0.2 | 0.7 | 2.0 | (0.3-12.0) | 0.4 |
| *Yersinia spp.* | 0 | 0.0 | 0 | 0.0 | | | | |
| *Rickettsia spp.* | 3 | 0.5 | 1 | 0.1 | 0.3 | 4.5 | (0.5-44.0) | 0.2 |
| **Viruses** | | | | | | | | |
| Adenovirus | 10 | 1.6 | 16 | 1.9 | 0.7 | 0.7 | (0.3-1.6) | 0.4 |
| Crimean-Congo hemorrhagic fever virus | 0 | 0.0 | 0 | 0.0 | | | | |
| Chikungunya virus | 3 | 0.5 | 1 | 0.1 | 0.3 | 4.5 | (0.5-44.0) | 0.2 |
| Cytomegalovirus | 16 | 2.5 | 16 | 1.9 | 0.5 | 1.3 | (0.7-2.7) | 0.4 |
| Dengue virus | 2 | 0.3 | 2 | 0.2 | 1.0 | 1.5 | (0.2-10.7) | 0.7 |
| Enterovirus | 0 | 0.0 | 0 | 0.0 | | | | |

| | | | | | | | | |
|---|---|---|---|---|---|---|---|---|
| Hepatitis E virus | 0 | 0.0 | 0 | 0.0 | | | | |
| Herpes Simplex Virus 1 | 0 | 0.0 | 0 | 0.0 | | | | |
| Herpes Simplex Virus 2 | 0 | 0.0 | 0 | 0.0 | | | | |
| Japanese encephalitis virus | 0 | 0.0 | 0 | 0.0 | | | | |
| Lassa A | 0 | 0.0 | 0 | 0.0 | | | | |
| Lassa B | 0 | 0.0 | 0 | 0.0 | | | | |
| Lassa virus (pan) | 2 | 0.3 | 3 | 0.4 | 1.0 | 1.0 | (0.2-6.1) | 1.0 |
| Measles virus | 1 | 0.2 | 1 | 0.1 | 1.0 | 1.3 | (0.1-21.3) | 0.8 |
| Mumps virus | 0 | 0.0 | 0 | 0.0 | | | | |
| Nipah virus | 0 | 0.0 | 1 | 0.1 | 1.0 | * | | |
| Parechovirus | 0 | 0.0 | 0 | 0.0 | | | | |
| Parvovirus B19 | 16 | 2.5 | 20 | 2.3 | 0.9 | 0.9 | (0.5-1.9) | 0.9 |
| Rubella virus | 8 | 1.2 | 19 | 2.2 | 0.2 | 0.7 | (0.3-1.5) | 0.3 |
| Rift valley fever virus | 0 | 0.0 | 0 | 0.0 | | | | |
| Varicella zoster virus | 1 | 0.2 | 1 | 0.1 | 1.0 | 1.3 | (0.1-21.3) | 0.8 |
| West Nile virus | 0 | 0.0 | 0 | 0.0 | | | | |
| Yellow fever virus | 0 | 0.0 | 0 | 0.0 | | | | |
| Zika virus | 6 | 0.9 | 3 | 0.4 | 0.2 | 2.7 | (0.7-10.8) | 0.2 |
| **Protozoa** | | | | | | | | |
| *Plasmodium falciparum* | 15 | 2.3 | 25 | 2.9 | 0.5 | 0.9 | (0.5-1.8) | 0.8 |
| *Plasmodium vivax* | 0 | 0.0 | 0 | 0.0 | | | | |
| *Toxoplasma gondii* | 0 | 0.0 | 0 | 0.0 | | | | |
| **Fungi** | | | | | | | | |
| *Candida albicans* | 3 | 0.5 | 6 | 0.7 | 0.7 | 0.6 | (0.2-2.5) | 0.5 |
| *Cryptococcus neoformans/gattii* | 0 | 0.0 | 0 | 0.0 | | | | |

* target detected only in controls ** target detected only in cases

Supplementary Table 16: Sensitivity analysis: Prevalence for each bacterial, viral, protozoal and fungal target, molecularly detected (PCR), and odds ratio for perinatal death, in maternal participants in cases and controls with good pregnancy outcomes, Kilifi County Hospital, Kenya.

| | Case N=318 | | Control N=315 | | Fisher's exact test | Odds Ratio | (95% CI) | p |
|---|---|---|---|---|---|---|---|---|
| | n | % | n | % | | | | |
| **Bacteria** | | | | | | | | |
| *Acinetobacter baumanii* | 0 | 0.0 | 0 | 0.0 | | | | |
| *Bartonella spp.* | 0 | 0.0 | 0 | 0.0 | | | | |
| *Brucella spp.* | 5 | 1.6 | 3 | 1.0 | 0.7 | 1.7 | (0.3-7.0) | 0.5 |
| *Burkholderia pseudommallei* | 0 | 0.0 | 0 | 0.0 | | | | |
| *Coxiella burnetii* | 1 | 0.3 | 0 | 0.0 | 1.0 | ** | | |
| *Escherichia coli/Shigella spp.* | 4 | 1.3 | 2 | 0.6 | 0.7 | 2.0 | (0.4-11.0) | 0.4 |
| *Enterococcus faecium* | 1 | 0.3 | 0 | 0.0 | 1.0 | ** | | |
| *Enterococcus faecalis* | 2 | 0.6 | 1 | 0.3 | 1.0 | 2.0 | (0.2-22.0) | 0.6 |
| group A streptococcus | 0 | 0.0 | 0 | 0.0 | | | | |
| group B streptococcus | 0 | 0.0 | 0 | 0.0 | | | | |
| *Haemophilus influenzae* type b | 0 | 0.0 | 0 | 0.0 | | | | |
| *Haemophilus influenzae* | 0 | 0.0 | 0 | 0.0 | | | | |
| *Klebsiella pneumoniae* | 0 | 0.0 | 0 | 0.0 | | | | |
| *Listeria monocytogenes* | 0 | 0.0 | 0 | 0.0 | | | | |
| *Leptospira spp* | 0 | 0.0 | 0 | 0.0 | | | | |
| *Moraxella catarrhalis* | 0 | 0.0 | 0 | 0.0 | | | | |
| *Mycobacterium tuberculosis* | 1 | 0.3 | 0 | 0.0 | 1.0 | ** | | |
| *Neisseria meningitidis* | 0 | 0.0 | 0 | 0.0 | | | | |
| *Neisseria gonorrhoeae* | 0 | 0.0 | 0 | 0.0 | | | | |
| *Orientia tsutsugamushi* | 0 | 0.0 | 0 | 0.0 | | | | |
| *Pseudomonas aeruginosa* | 2 | 0.6 | 0 | 0.0 | 0.5 | ** | | |
| *Streptococcus pneumoniae* | 1 | 0.3 | 1 | 0.3 | 1.0 | 1.0 | (0.1-15.9) | 1.0 |
| *Streptococcus suis* | 0 | 0.0 | 0 | 0.0 | | | | |
| *Salmonella enterica* serovar Paratyphi A | 1 | 0.3 | 0 | 0.0 | 1.0 | ** | | |
| *Salmonella spp.* | 1 | 0.3 | 0 | 0.0 | 1.0 | ** | | |
| *Salmonella enterica* serovar Typhi | 0 | 0.0 | 0 | 0.0 | | | | |
| *Staphylococcus aureus* | 1 | 0.3 | 0 | 0.0 | 1.0 | ** | | |
| *Treponema pallidum* | 1 | 0.3 | 1 | 0.3 | 1.0 | 1.0 | (0.1-15.9) | 1.0 |
| *Ureaplasma spp.* | 0 | 0.0 | 1 | 0.3 | 0.5 | * | | |
| *Yersinia spp.* | 0 | 0.0 | 0 | 0.0 | | | | |
| *Rickettsia spp.* | 3 | 0.9 | 1 | 0.3 | 0.6 | 3.0 | (0.3-29.9) | 0.3 |
| **Viruses** | | | | | | | | |
| Adenovirus | 3 | 0.9 | 1 | 0.3 | 0.6 | 3.0 | (0.3-30.0) | 0.3 |
| Crimean Congo Haemorrhagic Fever virus | 0 | 0.0 | 0 | 0.0 | | | | |
| Chikungunya virus | 3 | 0.9 | 1 | 0.3 | 0.6 | 3.0 | (0.3-29.0) | 0.3 |
| Cytomegalovirus | 8 | 2.5 | 7 | 2.2 | 1.0 | 1.1 | (0.4-3.2) | 0.8 |
| Dengue virus | 2 | 0.6 | 1 | 0.3 | 1.0 | 2.0 | (0.2-22.0) | 0.6 |
| Enterovirus | 0 | 0.0 | 0 | 0.0 | | | | |
| Hepatitis E virus | 0 | 0.0 | 0 | 0.0 | | | | |
| Herpes simplex virus 1 | 0 | 0.0 | 0 | 0.0 | | | | |

| | | | | | | | | |
|---|---|---|---|---|---|---|---|---|
| Herpes simplex virus 2 | 0 | 0.0 | 0 | 0.0 | | | | |
| Japanese encephalitis virus | 0 | 0.0 | 0 | 0.0 | | | | |
| Lassa A | 0 | 0.0 | 0 | 0.0 | | | | |
| Lassa B | 0 | 0.0 | 0 | 0.0 | | | | |
| Lassa virus (pan) | 2 | 0.6 | 3 | 1.0 | 0.7 | 0.7 | (0.1-4.0) | 0.7 |
| Measles virus | 0 | 0.0 | 1 | 0.3 | 0.5 | * | | |
| Mumps | 0 | 0.0 | 0 | 0.0 | | | | |
| Nipah virus | 0 | 0.0 | 1 | 0.3 | 0.5 | * | | |
| Parechovirus | 0 | 0.0 | 0 | 0.0 | | | | |
| Parvovirus B19 | 4 | 1.3 | 4 | 1.3 | 1.0 | 1.0 | (0.3-4.0) | 1.0 |
| Rubella virus | 8 | 2.5 | 13 | 4.1 | 0.3 | 0.6 | (0.2-1.5) | 0.3 |
| Rift valley fever virus | 0 | 0.0 | 0 | 0.0 | | | | |
| Varicella zoster virus | 1 | 0.3 | 0 | 0.0 | 1.0 | ** | | |
| West Nile virus | 0 | 0.0 | 0 | 0.0 | | | | |
| Yellow fever virus | 0 | 0.0 | 0 | 0.0 | | | | |
| Zika virus | 2 | 0.6 | 2 | 0.6 | 1.0 | 1.0 | (0.1-7.1) | 1.0 |
| **Protozoa** | | | | | | | | |
| *Plasmodium falciparum* | 13 | 4.1 | 14 | 4.4 | 0.8 | 0.9 | (0.4-2.0) | 0.8 |
| *Plasmodium vivax* | 0 | 0.0 | 0 | 0.0 | | | | |
| *Toxoplasma gondii* | 0 | 0.0 | 0 | 0.0 | | | | |
| **Fungi** | | | | | | | | |
| *Candida albicans* | 0 | 0.0 | 2 | 0.6 | 0.2 | * | | |
| *Cryptococcus neoformans/gattii* | 0 | 0.0 | 0 | 0.0 | | | | |

* target detected only in controls ** target detected only in cases

Supplementary Table 17: Prevalence of bacterial targets molecularly detected (PCR) in blood, and bacteraemia (conventional culture), by species, and odds ratio for perinatal death, Kilifi County Hospital, Kenya and Hiwot Fana Comprehensive Specialised Hospital, Ethiopia.

| | **Case N=642** | | **Control N=855** | | **Odds Ratio** | **(95% CI)** | **p** |
|---|---|---|---|---|---|---|---|
| | **n** | **%** | **n** | **%** | | | |
| *Escherichia coli/Shigella spp.* | 18 | 2.8 | 10 | 1.2 | 2.2 | (1.0-4.8) | 0.050 |
| group A streptococcus | 2 | 0.3 | 0 | 0.0 | ** | | |
| group B streptococcus | 3 | 0.5 | 1 | 0.1 | 3.3 | (0.4-32.3) | 0.3 |
| *Neisseria gonorrhoeae* | 2 | 0.3 | 2 | 0.2 | 1.2 | (0.2-8.6) | 0.9 |
| *Salmonella enterica* serovar Paratyphi A | 2 | 0.3 | 2 | 0.2 | 1.2 | (0.2-8.6) | 0.9 |
| *Salmonella spp.* | 4 | 0.6 | 3 | 0.4 | 1.6 | (0.3-7.0) | 0.6 |
| *Staphylococcus aureus* | 7 | 1.1 | 5 | 0.6 | 1.6 | (0.5-5.1) | 0.4 |

* target detected/bacteraemia only in controls ** target detected/bacteraemia only in cases

Supplementary Table 18: Prevalence of each bacterial, viral, fungal, parasitic target molecularly detected (PCR) in vagino-rectal swabs, by species, and odds ratio for perinatal death, Hiwot Fana Comprehensive Specialised Hospital, Ethiopia.

| | Case N=324 | | Control N=347 | | Odds Ratio | (95% CI) | p |
|---|---|---|---|---|---|---|---|
| **Bacteria** | **n** | **%** | **n** | **%** | | | |
| *Escherichia coli/Shigella spp.* | 140 | 43 | 111 | 32 | 1.6 | (1.2-2.2) | 0.003 |
| group A streptococcus | 7 | 2 | 3 | 0.9 | 2.5 | (0.7-9.9) | 0.2 |
| group B streptococcus | 58 | 18 | 52 | 15 | 1.2 | (0.8-1.9) | 0.3 |
| *Haemophilus influenzae* type b | 0 | 0 | 0 | 0 | | | |
| *Haemophilus influenzae* | 1 | 0 | 5 | 1.4 | 0.2 | (0.0-1.8) | 0.2 |
| *Klebsiella pneumoniae* | 28 | 9 | 24 | 6.9 | 1.3 | (0.7-2.3) | 0.4 |
| *Listeria monocytogenes* | 0 | 0 | 1 | 0.3 | * | | |
| *Neisseria meningitidis* | 8 | 2 | 12 | 3.5 | 0.7 | (0.3-1.8) | 0.5 |
| *Orientia tsutsugamushi* | 0 | 0 | 0 | 0 | | | |
| *Pseudomonas aeruginosa* | 13 | 4 | 14 | 4 | 1.0 | (0.4-2.2) | 1.0 |
| *Streptococcus pneumoniae* | 39 | 12 | 39 | 11 | 1.1 | (0.7-1.7) | 0.7 |
| *Salmonella enterica* serovar Paratyphi A | 1 | 0 | 1 | 0.3 | 1.1 | (0.1-17.2) | 1.0 |
| *Salmonella spp.* | 1 | 0 | 2 | 0.6 | 0.5 | (0.1-5.9) | 0.6 |
| *Salmonella enterica* serovar Typhi | 2 | 1 | 2 | 0.6 | 1.1 | (0.2-7.7) | 0.9 |
| *Staphylococcus aureus* | 38 | 12 | 44 | 13 | 0.9 | (0.6-1.5) | 0.7 |
| *Treponema pallidum* | 3 | 1 | 3 | 0.9 | 1.1 | (0.2-5.4) | 0.9 |
| *Yersinia spp.* | 0 | 0 | 0 | 0 | | | |
| *Rickettsia spp.* | 0 | 0 | 0 | 0 | | | |
| **Viruses** | | | | | | | |
| Adenovirus | 8 | 2 | 11 | 3.2 | 0.8 | (0.3-2.0) | 0.6 |
| Chikungunya virus | 0 | 0 | 0 | 0 | | | |
| Dengue virus | 0 | 0 | 1 | 0.3 | * | | |
| Enterovirus | 0 | 0 | 0 | 0 | | | |
| Parechovirus | 0 | 0 | 0 | 0 | | | |
| Rubella virus | 0 | 0 | 0 | 0 | | | |
| Zika virus | 1 | 0 | 3 | 0.9 | 0.4 | (0.0-3.4) | 0.4 |
| **Protozoa** | | | | | | | |
| *Plasmodium falciparum* | 0 | 0 | 0 | 0 | | | |
| *Plasmodium vivax* | 0 | 0 | 0 | 0 | | | |
| *Toxoplasma gondii* | 1 | 0 | 1 | 0.3 | 1.1 | (0.1-17.2) | 1.0 |
| **Fungi** | | | | | | | |
| *Cryptococcus neoformans/gattii* | 0 | 0 | 0 | 0 | | | |

* target detected only in controls ** target detected only in cases

Supplementary Table 19: Prevalence of each bacterial species conventionally cultured from vagino-rectal swabs, and odds ratio for perinatal death, Hiwot Fana Comprehensive Specialised Hospital, Ethiopia.

| | **Case N=324** | | **Control N==347** | | **Odds Ratio** | **(95% CI)** | **p** |
|---|---|---|---|---|---|---|---|
| | **n** | **%** | **n** | **%** | | | |
| **Bacteria** | | | | | | | |
| *Citrobacter spp.* | 1 | 0.3 | 4 | 1.2 | 0.3 | (0.0-2.4) | 0.2 |
| *Escherichia coli/Shigella spp.* | 193 | 59.6 | 199 | 57.3 | 1.1 | (0.8-1.5) | 0.6 |
| *Enterobacter spp.* | 8 | 2.5 | 11 | 3.2 | 0.8 | (0.3-2.0) | 0.6 |
| *Enterococcus faecalis* | 7 | 2.2 | 4 | 1.2 | 1.9 | (0.6-6.5) | 0.3 |
| group B streptococcus | 26 | 8.0 | 19 | 5.5 | 1.5 | (0.8-2.8) | 0.2 |
| group C/G streptococcus | 7 | 2.2 | 4 | 1.2 | 1.7 | (0.7-4.5) | 0.3 |
| group D streptococcus | 145 | 44.8 | 156 | 45.0 | 1.0 | (0.7-1.3) | 1.0 |
| *Klebsiella pneumoniae* | 25 | 7.7 | 20 | 5.8 | 1.4 | (0.7-2.5) | 0.3 |
| *Proteus sp.* | 2 | 0.6 | 0 | 0.0 | ** | | |
| *Pseudomonas fluorescens* | 1 | 0.3 | 0 | 0.0 | ** | | |
| **Fungi** | | | | | | | |
| *Candida albicans* | 4 | 1.2 | 5 | 1.4 | 0.9 | (0.2-3.2) | 0.8 |

* target detected only in controls ** target detected only in cases

Supplementary Table 20: Odds ratios for association of *Escherichia Coli* on vagino-rectal swab molecularly detected (PCR) with perinatal death in Hiwot Fana Comprehensive Specialised Hospital, Ethiopia, univariable and multivariable analyses.

| | **Univariable analyses** | | | **Multivariable analyses** | | |
|---|---|---|---|---|---|---|
| | **OR** | **95%CI** | **p** | **OR** | **95%CI** | **p** |
| ***Escherichia Coli*** | | | | | | |
| No | 1 | | | 1 | | |
| Yes | 1.62 | (1.18-2.22) | 0.003 | 1.31 | (0.91-2.90) | 0.15 |
| **Age** | | | | | | |
| <20 | 1.45 | (0.95-2.50) | | 1.27 | (0.91-1.90) | |
| 20 to <30 | 1 | | | 1 | | |
| 30 to <40 | 1.88 | (1.30-2.73) | | 1.54 | (0.69-2.31) | |
| ≥40years | 0.66 | (0.22-1.97) | 0.004 | 0.40 | (0.13-1.25) | 0.048 |
| **Marital status** | | | | | | |
| Married | 1 | | | | | |
| Single | 1.05 | (0.07-16.90) | | | | |
| Divorced | * | | | | | |
| Widowed | * | | 1 | | | |
| **Education level** | | | | | | |
| None | 3.88 | (2.51-6.00) | | 3.74 | (2.31-6.06) | |
| Primary | 1 | | | 1 | | |
| Secondary | 0.75 | (0.38-1.48) | | 0.70 | (0.33-1.48) | |
| Higher | 1.6 | (0.84-3.03) | <0.001 | 1.70 | (0.84-3.45) | <0.001 |
| **Nulliparous** | | | | | | |
| No | 1 | | | | | |
| Yes | 0.96 | (0.68-1.35) | 0.8 | | | |
| **Sex** | | | | | | |
| Male | 1.18 | (0.85-1.62) | 0.3 | 1.00 | (0.69-1.44) | |
| Female | 1 | | | 1 | | 1 |

*insufficient data

Supplementary Table 21: Prevalence of each bacterial, viral, and fungal target molecularly detected (PCR) in oropharyngeal swabs, by species, and odds ratio for perinatal death, Hiwot Fana Comprehensive Specialised Hospital, Ethiopia.

| | **Case N=324** | | **Control N=327** | | **OR** | **95%CI** | **p** |
|---|---|---|---|---|---|---|---|
| | **n** | **%** | **n** | **%** | | | |
| **Bacteria** | | | | | | | |
| *Acinetobacter baumanii* | 15 | 4.6 | 19 | 5.5 | 0.8 | (0.4-1.7) | 0.6 |
| *Bordatella sp.* | 9 | 2.8 | 2 | 0.6 | 1.9 | (1.1-23.0) | 0.042 |
| *Brevimundus vesicularis* | 0 | 0 | 0 | 0.0 | | | |
| *Burkholderia pseudommallei* | 1 | 0.3 | 0 | 0.0 | ** | | |
| *Corynebacterium. diphtheriae* | 0 | 0 | 0 | 0.0 | | | |
| *Corynebacterium pneumoniae* | 0 | 0 | 0 | 0.0 | | | |
| *Corynebacterium ulcerans/pseudotb* | 2 | 0.6 | 1 | 0.3 | 2.2 | (0.2-23.8) | 0.5 |
| *Chlamydia trachomatis* | 0 | 0 | 0 | 0.0 | | | |
| DT (toxin) | 0 | 0 | 0 | 0.0 | | | |
| *Escherichia coli/Shigella spp.* | 0 | 0 | 0 | 0.0 | | | |
| group A streptococcus | 3 | 0.9 | 0 | 0.0 | ** | | |
| group B streptococcus | 16 | 4.9 | 10 | 2.9 | 1.8 | (0.8-3.9) | 0.3 |
| *Haemophilus influenzae* type b | 10 | 3.1 | 5 | 1.4 | 2.2 | (0.7-6.4) | 0.2 |
| *Haemophilus influenzae* | 19 | 5.9 | 22 | 6.3 | 0.9 | (0.5-1.7) | 0.8 |
| *Klebsiella pneumoniae* | 8 | 2.5 | 10 | 2.9 | 0.9 | (0.3-2.2) | 0.7 |
| *Listeria monocytogenes* | 0 | 0 | 0 | 0.0 | | | |
| *Moraxella catarrhalis* | 14 | 4.3 | 15 | 4.3 | 1.0 | (0.5-2.1) | 1.0 |
| *Mycobacterium tuberculosis* | 2 | 0.6 | 1 | 0.3 | 2.2 | (0.2-23.8) | 0.5 |
| *Mycoplasma pneumoniae* | 0 | 0 | 0 | 0.0 | | | |
| *Pseudomonas aeruginosa* | 10 | 3.1 | 3 | 0.9 | 1.3 | (0.9-1.8) | 0.2 |
| *Streptococcus pneumoniae* | 94 | 29 | 85 | 24.5 | 1.3 | (0.9-1.8) | 0.2 |
| *Staphylococcus aureus* | 31 | 9.6 | 28 | 8.1 | 1.2 | (0.7-2.1) | 0.5 |
| *Ureaplasma spp.* | 4 | 1.2 | 7 | 2.0 | 0.6 | (0.2-2.1) | 0.4 |
| **Viruses** | | | | | | | |
| Adenovirus | 2 | 0.6 | 4 | 1.2 | 0.5 | (0.1-2.9) | 0.5 |
| hCoV 229E | 0 | 0 | 2 | 0.6 | * | | |
| hCoV HKU1 | 1 | 0.3 | 2 | 0.6 | 0.5 | (0.1-5.9) | 0.6 |
| hCoV NL63 | 2 | 0.6 | 1 | 0.3 | 2.2 | (0.2-23.8) | 0.5 |
| hCoV OC43 | 5 | 1.5 | 5 | 1.4 | 1.1 | (0.3-3.7) | 0.9 |
| Cytomegalovirus | 1 | 0.3 | 4 | 1.2 | 0.3 | (0.0-2.4) | 0.2 |
| Enterovirus | 3 | 0.9 | 8 | 2.3 | 0.4 | (0.1-1.5) | 0.2 |
| Human metapneumovirus | 0 | 0 | 1 | 0.3 | * | | |
| Influenza A | 0 | 0 | 1 | 0.3 | * | | |
| Influenza B | 0 | 0 | 0 | 0.0 | | | |
| Measles virus | 0 | 0 | 0 | 0.0 | | | |
| MERS Coronavirus N | 4 | 1.2 | 2 | 0.6 | 2.2 | (0.4-11.9) | 0.4 |
| MERS-CoV upE | 0 | 0 | 0 | 0.0 | | | |
| Parainfluenza 1 | 0 | 0 | 0 | 0.0 | | | |
| Parainfluenza 2 | 0 | 0 | 0 | 0.0 | | | |
| Parainfluenza 3 | 1 | 0.3 | 0 | 0.0 | ** | | |

| | | | | | | | |
|---|---|---|---|---|---|---|---|
| Parainfluenza 4 | 0 | 0 | 0 | 0.0 | | | |
| Parvovirus B19 | 0 | 0 | 1 | 0.3 | * | | |
| Rubella virus | 0 | 0 | 0 | 0.0 | | | |
| Respiratory syncytial virus | 0 | 0 | 0 | 0.0 | | | |
| Rhinovirus | 1 | 0.3 | 7 | 2.0 | 0.2 | (0.0-1.2) | 0.077 |
| Varicella zoster virus | 0 | 0 | 0 | 0.0 | | | |
| **Fungi** | | | | | | | |
| *P. jirovecii* | 0 | 0 | 0 | 0.0 | | | |

* target detected only in controls ** target detected only in cases

Supplementary Table 22 Odds ratios for association of *Bordatella spp.* on oropharyngeal swab (PCR) with perinatal death in Hiwot Fana Comprehensive Specialised Hospital, Ethiopia; univariable and multivariable analyses

| | Univariable analyses | | | Multivariable analyses | | |
|---|---|---|---|---|---|---|
| | **OR** | **95%CI** | **p** | **OR** | **95%CI** | **p** |
| ***Bordatella spp.*** | | | | | | |
| No | 1 | | | | | |
| Yes | **4.93** | **(1.06-22.98)** | | **all are cases*** | | |
| **Age** | | | | | | |
| <20 | 1.45 | (0.95-2.50) | | 1.21 | (0.66-2.23) | |
| 20 to <30 | 1 | | | 1 | | |
| 30 to <40 | 1.88 | (1.30-2.73) | | 1.52 | (1.00-2.32) | |
| ≥40years | 0.66 | (0.22-1.97) | 0.004 | 0.33 | (0.10-1.10) | 0.035 |
| **Marital status** | | | | | | |
| Married | 1 | | | | | |
| Single | 1.05 | (0.07-16.90) | | | | |
| Divorced | ** | | | | | |
| Widowed | ** | | 1 | | | |
| **Education level** | | | | | | |
| None | 3.88 | (2.51-6.00) | | 4.00 | (2.45-6.52) | |
| Primary | 1 | (0.38-1.48) | | 1 | | |
| Secondary | 0.75 | (0.38-1.48) | | 0.66 | (0.31-1.42) | |
| Higher | 1.6 | (0.84-3.03) | <0.001 | 1.79 | (0.88-3.64) | <0.001 |
| **Nulliparous** | | | | | | |
| No | 1 | | | | | |
| Yes | 0.96 | (0.68-1.35) | 0.8 | | | |
| **Sex baby/fetus** | | | | | | |
| Male | 1.18 | (0.85-1.62) | 0.3 | 0.98 | (0.67-1.41) | 0.9 |
| Female | 1 | | | 1 | | |

*not included in the model as all *Bordatella spp.* were in cases

**insufficient data

Supplementary Figure 1: Prevalence of bacterial species, detected by PCR in blood in maternal participants in Kilifi County Hospital, Kenya, for cases, all controls and a subset of controls with good pregnancy outcomes.*

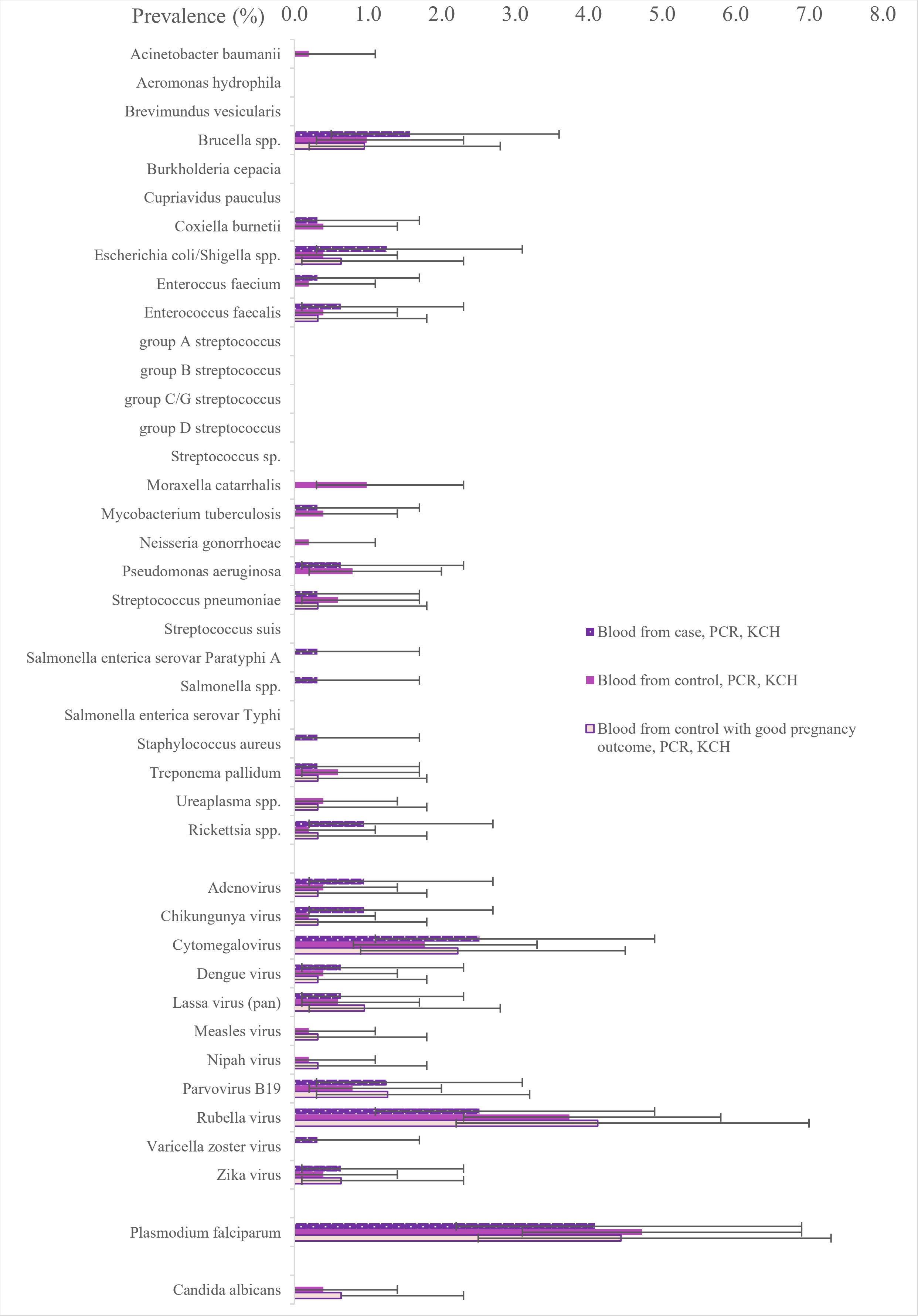

**Error bars show 95% confidence intervals
